# Whole Exome Sequencing Uncovers Key Genetic Variants in Congenital Tooth Agenesis: An Integrative Omics Approach

**DOI:** 10.1101/2024.11.26.24317461

**Authors:** Prashant Ranjan, Chandra Devi, Neha Verma, Rajesh Bansal, Vinay Kumar Srivastava, Parimal Das

**Affiliations:** Centre for Genetic Disorders, Institute of Science, Banaras Hindu University, Varanasi, BHU-221005, UP, India; Dentistry Oral Surgery and Medicine, Institute of Medical Sciences, Banaras Hindu University, Varanasi, BHU-221005, UP, India; Dr Bhimrao Ramji Ambedkar Government Medical College, Kannauj, UP, India

**Keywords:** Omics integration, Diseases association, Compound heterozygous, Bioinformatics, Molecular Dynamics

## Abstract

This study investigates the genetic underpinnings of congenital tooth agenesis (CTA) using a multi-omics approach, integrating whole exome sequencing (WES)and RNA expression analysis. WES was used to analyze the genetic basis of CTA in six affected individuals one with syndromic and five with non-syndromic CTA alongside three healthy and two internal controls. We identified both known and novel variants in candidate genes (*EDA, WNT10A, PAX9, TSPEAR*) and assessed the functional impacts of novel variants (WNT10A (A135S), and compound heterozygous *TSPEAR (*L219P, I419Lfs*150) using RT-PCR, while bioinformatics tools were applied to both known and novel variants. RT-PCR indicated disrupted EDA and WNT10A signaling in novel candidate genes *WNT10A* and *TSPEAR*. Computational analysis showed deleterious effects for six variants, with gene ontology, protein disorder, localization, and post-translational modifications suggesting significant functional changes. Molecular dynamics simulations predicted that these variants could impact protein stability and function. Additionally, WES analysis revealed 21 genes consistently present in all patients (MAF ≤20%), including novel variants in *OR4F21* (K310R, F44L) and *LCORL* (L1734P). Two variants, *OR4F21* (K310R) and *MRTFB* (A135A), appeared in all cases. Furthermore, 391 genes were shared among three patients, 204 among four, and 98 among five. Integrating multi-omic data from the GEO database identified 18 upregulated and 15 downregulated genes, with variants linked to systemic conditions such as autism, Alzheimer’s, congenital heart disease (CHD), ALS (amyotrophic lateral sclerosis) and cancer.

Our findings provide insights into CTA’s molecular mechanisms, identifying potential biomarkers and therapeutic targets. Further validation could improve diagnosis and treatment strategies for CTA.

**Graphical Abstract:** 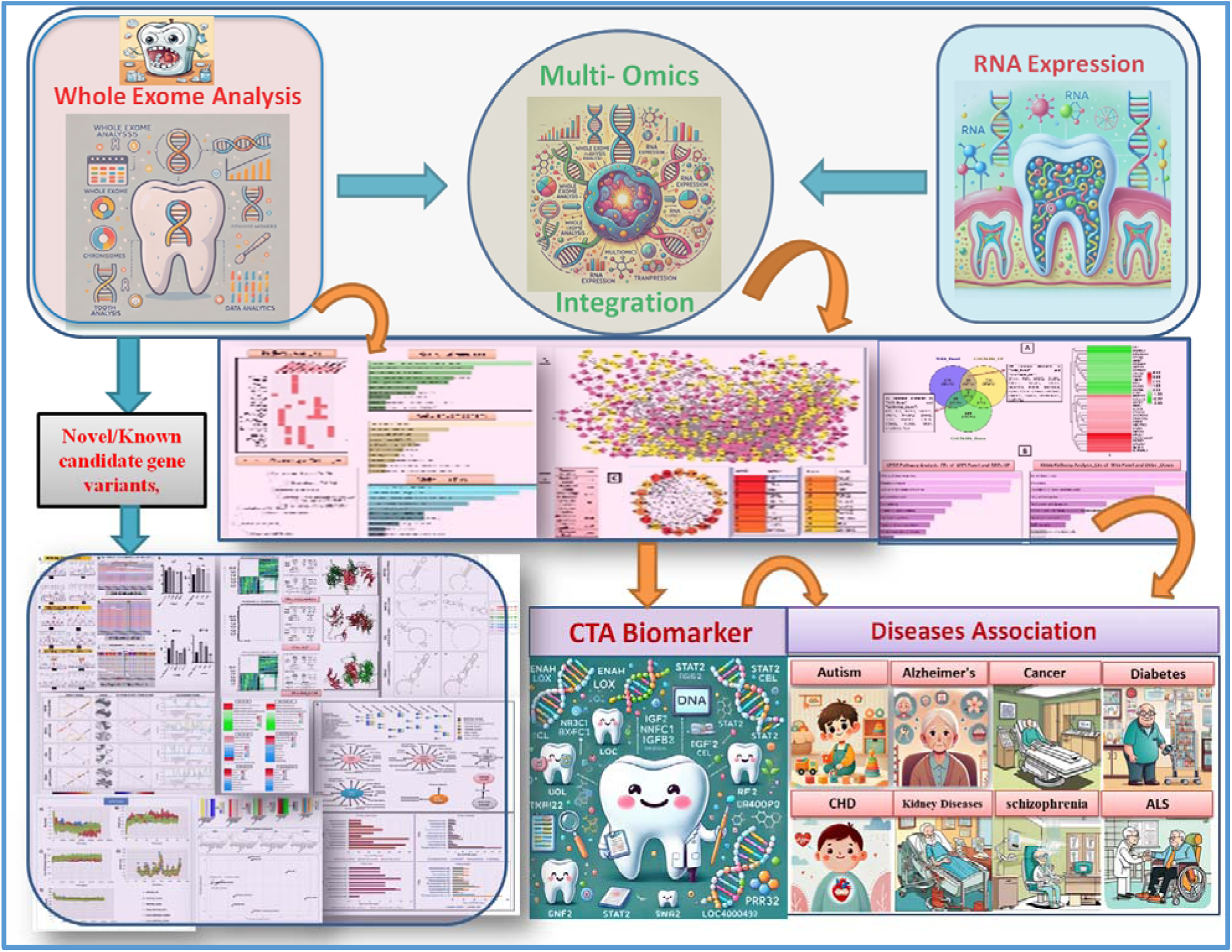

## 1. Introduction

Congenital tooth agenesis (CTA) is a developmental anomaly marked by the absence of one or more teeth, making it one of the most prevalent dental irregularities observed in humans. CTA is categorized based on the number of missing teeth: hypodontia involves the absence of one to five teeth (excluding third molars), oligodontia refers to the absence of six or more teeth, and anodontia denotes the complete absence of all teeth, which is extremely uncommon (Schonberger et al., 2023). CTA may present as a non-syndromic condition, occurring independently, or as a component of a syndromic disorder that also affects other ectodermal tissues, such as hair, nails, and sweat glands (Meade & Dreyer, 2023).

Non-syndromic congenital tooth agenesis (CTA) is estimated to affect around 8% of the global population, though prevalence rates vary widely by region. For example, some European studies have reported rates as low as 0.4%, reflecting the complex genetic factors influencing this condition(Galluccio et al., 2012). Inheritance patterns for non-syndromic CTA include autosomal dominant, autosomal recessive, and X-linked modes, highlighting its genetic heterogeneity (Yu et al., 2019).

Over the past few decades, various genes have been implicated in CTA. Early research identified mutations in genes such as *MSX1* and *PAX9*, both essential for tooth development, as key contributors to non-syndromic CTA. These genes are expressed in the dental mesenchyme and play a role in regulating other critical genes like BMP4, which is necessary for odontogenesis (Galluccio et al., 2012). Additionally, genes in the Wnt signaling pathway, including *WNT10A* and *AXIN2*, have been linked to CTA, with *AXIN2* mutations particularly associated with familial cases of oligodontia (Yu et al., 2019). The X-linked gene *EDA*, which is crucial for ectodermal development, has also been connected to both syndromic and non-syndromic tooth agenesis(Gao et al., 2023).

In our study, whole exome sequencing (WES) was employed to explore the genetic underpinnings of CTA. We analyzed exome data from six affected individuals (five with non-syndromic CTA and one with syndromic CTA), as well as three matched healthy controls and two internal family controls. This analysis revealed both previously reported and novel variants in candidate genes, including *EDA*, *WNT10A*, *PAX9*, and *TSPEAR*. Bioinformatics analysis were conducted to assess the potential functional impact of these variations. Our WES data identified 21 genes consistently present across all six patients, filtered based on a minor allele frequency (MAF) of ≤20%. We also observed 391 genes shared among three patients, 204 among four patients, and 98 genes shared by five patients, which enabled us to refine a gene panel focused on genes identified in at least three CTA samples. To enhance our genetic findings, we integrated multi-omic data from the Gene Expression Omnibus (GEO) database, specifically examining syndromic CTA cases. This integrated approach revealed 18 upregulated and 15 downregulated genes from our WES panel. Our analysis identified three novel variants associated with tooth agenesis, along with several other gene variants previously unlinked to CTA but known to play roles in other conditions. These findings underscore the significant genetic contributions to congenital tooth agenesis, offering new insights that could improve diagnostic and therapeutic approaches. Enhanced genetic understanding may facilitate early diagnosis, enabling timely, targeted interventions that can improve outcomes for those affected by CTA.

## 2. Methodology

### 2.1 Study Participants and Ethical Approval

The Declaration of Helsinki’s ethical standards were followed in this investigation. All participants or, in the case of minors, their parents or legal guardians, provided written informed consent. The Institute of Science’s Ethics Committee of Banaras Hindu University in India examined and approved the research procedure. CTA patients were among the participants. Participation conditions were a verified diagnosis of CTA, being between the ages of 18 and 35, having complete dental records and radiographic images available, and having written consent from participants or their legal guardians. The exclusion criteria included a history of orthodontic treatment, inadequate dental records, or tooth loss due to non-congenital causes. The Institute of Medical Sciences, India’s Department of Dentistry, Oral Surgery, and Medicine provided the five control samples and six patients for the study. The patients were booked for dental implant treatments after undergoing clinical evaluations. Five control subjects, who were all between the ages of 20 and 40, were chosen in order to closely resemble the patients’ demographics. To maintain confidentiality and anonymity, each sample was assigned a unique study code.

### 2.2 Data Collection

Dental records and radiographs of each participant were carefully examined to confirm the diagnosis and determine the extent of CTA. Clinical assessments were performed by qualified dental professionals to ensure the accuracy and consistency of data collection.

### 2.3 Blood Collection, Storage, and DNA/RNA Isolation

Peripheral blood samples (1.5 mL) were collected using a 5 mL heparinized syringe and transported to the laboratory within 60 minutes, stored in a cooling box to preserve sample integrity. Genomic DNA was first extracted from these samples using the standardized salting-out method. The quality and quantity of the DNA were evaluated through spectrophotometric analysis (NanoDrop 2000, Thermo Scientific Inc.) or fluorometry-based methods, including the DNA Assay BR (Invitrogen, Cat# Q32853) and the Qubit High Sensitivity Assay (Invitrogen). For RNA isolation, 200 μL of blood was processed using the Trizol reagent (Sigma-Aldrich, Catalog No. T9424). The concentration and purity of the extracted RNA were assessed using a NanoDrop spectrophotometer (Thermo Scientific, USA).

### 2.4 WES Analysis

WES was conducted utilizing the Illumina Next-Generation Sequencing platform, focusing on approximately 30Mb of the human exome. This approach encompassed nearly 99% of the coding regions as defined by CCDS and RefSeq. Sequencing achieved a mean depth of 80-100X, with over 90% of the targeted regions covered at a minimum depth of 20X. To ensure data quality, duplicate reads were eliminated, and base quality recalibration was performed. The reads were mapped to the human reference genome GRCh38. Variant calling followed the GATK best practices, and subsequent variant annotation was carried out using databases such as OMIM, GWAS, gnomAD, and 1000 Genomes. Filtration of the annotated variants was based on several criteria, including a gnomAD allele frequency ≤ 0.01 or unavailable, a minimum read depth of 20, pathogenicity, and predicted functional consequences. Upon confirming the presence of the mutant variant in the candidate gene with a minor allele frequency (MAF) of ≤ 0.01.

The WES analysis was conducted on a small Indian cohort consisting of 11 samples: 6 patient samples (DEN1, DEN10, DEN12, DEN20.1, DEN24.1, DEN23.1) and 5 controls, which included 3 healthy individuals and 2 internal controls. Notably, samples DEN1 and DEN12 were sourced from (Ranjan Jr et al., 2024; Sarkar et al., 2014). Initially, variants were filtered across all samples using a Minor Allele Frequency (MAF) threshold of ≤0.20 to focus on rare variants that may be relevant to tooth agenesis. Following this, variants detected in control samples were subtracted from those found in the patient data, thus removing common variants likely unrelated to the condition.

Subsequent analysis involved identifying genes shared among different numbers of patients by examining the presence of variants across 3, 4, 5, or all 6 patient samples. This was accomplished using a combination of Excel and Python scripting to facilitate accurate comparison across datasets. Genes that appeared in at least 3 out of the 6 patient samples were selected to form the final WES gene panel, prioritizing those with a higher likelihood of being associated with tooth agenesis based on their recurrence across multiple patients.

### 2.5 Sanger Sequencing

The salting-out technique was used to separate DNA from patient samples in order to verify new TSPEAR and WNT10A variations. Using certain primers, the TSPEAR and WNT10A sections were amplified by PCR and purified. Particular primers surrounding the target areas in TSPEAR and WNT10A were created and verified for the ideal melting temperature in order to perform Sanger sequencing. Initial denaturation at 95°C for 5 minutes, 30–35 cycles of 95°C for 30 seconds, primer-specific annealing at 55–60°C for 30 seconds, extension at 72°C for 45–60 seconds, and final extension at 72°C for 5 minutes were the processes used in PCR amplification. Following confirmation of the anticipated size on a 1.5% agarose gel, PCR products were purified to get rid of extra primers and nucleotides. The purified PCR products were subjected to Sanger sequencing using a sequencing dye terminator mix. Sequencing reactions included initial denaturation at 96°C for 1 minute, followed by 25–30 cycles of 96°C for 10 seconds, annealing at 50–55°C for 5 seconds, and extension at 60°C for 4 minutes. Sequencing reaction products were then purified and loaded onto a capillary electrophoresis instrument for analysis. Chromatograms were analyzed to confirm the presence of nucleotide changes, with mutant changes indicated by red arrows and unchanged nucleotides by green arrows Figure2.

### 2.6 RT PCR

Blood samples from patients were used to extract total RNA, which was then measured using spectrophotometry to guarantee purity. Using oligo(dT) primers and the RevertAid First Strand cDNA Synthesis Kit (Thermo Scientific, Catalog #K1622, USA), reverse transcription of mRNA was carried out in accordance with the manufacturer’s instructions. The QuantStudio 6 Flex Real-Time PCR System from Applied Biosystems (Thermo Scientific, USA) was used to perform quantitative PCR (qRT-PCR). One microliter of cDNA (diluted 1:5), 6.25 microliters of Maxima SYBR Green/ROX qPCR Master Mix (2X, Thermo Scientific, Catalog #K0221, USA), one microliter of 10 μM forward primer, one microliter of 10 μM reverse primer specific for EDA or WNT10A, and nuclease-free water were all included in the 12.5 microliter reaction mixture. Following two minutes of initial denaturation at 95°C, 40 cycles of 95°C for 15 seconds and 60°C for 30 seconds comprised the qPCR cycling conditions. Primer blast utilized Primer designing. The ΔCT technique was used to quantify relative expression levels after normalizing gene expression to an endogenous control (Beta-Actin).

### 2.7 Statistical Analysis

GraphPad Prism software (version 8; GraphPad Software, La Jolla, CA, USA) was used to conduct statistical analyses (Swift, 1997). To identify significant differences between groups, a one-way analysis of variance (ANOVA) was employed. To find particular group differences, post hoc comparisons were carried out using Tukey’s multiple comparison test, with a significance threshold set at p < 0.05. The mean ± standard deviation (SD) of the data was displayed.

### 2.8 Multiomics Integration

#### 2.8.1 Data Sources and Gene Selection

Differentially expressed genes (DEGs) were obtained from the study by(Ranjan Jr et al., 2024), using the GEO dataset ID **GSE56486**. This dataset was analyzed to identify genes that were significantly upregulated or downregulated. WES data was also utilized, comprising a gene panel with 709 genes relevant to congenital tooth agenesis.

#### 2.8.2 Identification of Common Genes

To identify common genetic elements, the list of 709 WES genes was compared against the list of DEGs from the GSE56486 dataset. The comparison was performed using **Venny 2.0** (Oliveros, 2007), a widely-used tool for visualizing and identifying overlapping genes between datasets. Common genes between the WES panel and both upregulated and downregulated DEGs were identified, providing a focused set of targets for further analysis.

#### 2.8.3 Enrichment Analysis of WES Panel

WES data was analyzed to identify genetic variants associated with congenital tooth agenesis. Variants from the WES panel were annotated and filtered based on established criteria, including allele frequency, functional impact predictions, and known associations with dental development. Following variant filtration, enrichment analysis was performed using Enrichr(E. Y. Chen et al., 2013), a comprehensive tool that allows for the functional characterization of gene lists. The analysis included pathways, biological processes, and disease ontology terms to determine if specific molecular pathways or biological functions were significantly overrepresented among the identified genes.

#### 2.8.4 Enrichment Analysis of Common Genes in DEGs

Microarray-seq data analysis was used to identify differentially expressed genes (DEGs). To identify genes shared by the two datasets, genes exhibiting a significant differential expression (p-value < 0.05) were compared to the WES panel. After that, Enrichr was used to do an enrichment analysis on the shared genes (E. Y. Chen et al., 2013)(E. Y. This method assisted in the identification of cellular components, molecular processes, and enriched biological pathways linked to the genes that exhibit both genetic variation and differential expression. The research offered a more thorough comprehension of the molecular pathways behind congenital tooth agenesis by combining the information from WES and DEGs.

#### 2.8.5 PPI Network Construction and Analysis

A list of 709 genes was examined using the STRING database (Szklarczyk et al., 2010) in order to look into the protein-protein interaction (PPI) network among the genes from the WES panel. The interaction score threshold was set to a high confidence level (≥0.4) to ensure dependability when the genes were submitted to STRING in order to find known and anticipated interactions. Finding possible clusters and functional relationships within the network that could be connected to congenital tooth agenesis was the goal of this investigation. Then, using Cytoscape (Doncheva et al., 2018) a powerful tool for network visualization and analysis, the PPI network data from STRING was exported and loaded. After the network was further optimized within Cytoscape, interconnecting clusters and paths could be seen more clearly. The analysis included the calculation of topological parameters such as node degree, betweenness centrality, and clustering coefficients, which are essential for identifying key characteristics of the network structure.

#### 2.8.6 Hub Gene Identification

To pinpoint central genes playing crucial roles within the PPI network, hub gene analysis was performed using **CytoHubba** (Chin et al., 2014), a plugin for Cytoscape. CytoHubba offers multiple algorithms to rank nodes based on their importance, using metrics such as **Degree**, **Betweenness**, **Closeness**, and **EPC (Edge Percolated Component)** scores. By applying these algorithms, the genes were ranked, and those with the highest centrality measures were identified as hub genes, indicating their potential key regulatory roles in the network.

The hub genes were visually highlighted within the PPI network, and sub-networks were extracted to focus on significant interactions involving these central nodes. This approach enabled the identification of core molecular interactions and pathways, providing deeper insights into the biological mechanisms that may contribute to congenital tooth agenesis.

### 2.9 Pathogenicity Prediction

To predict the pathogenic potential of mutant proteins, we utilized a range of algorithm-based online tools, including SIFT (Vaser et al., 2016), PolyPhen-2 (Adzhubei & Jordan, n.d.), MutationTaster (Schwarz et al., 2010) and CADD (Rentzsch et al., 2019). Each tool required mutation-specific input, such as details of the amino acid substitution, and provided predictions based on distinct threshold and cutoff values.

### 2.10 Protein Stability Prediction

We used a number of methods based on Gibbs free energy estimates to investigate how mutations affect protein stability. The stability changes linked to the mutant proteins were predicted using tools like mCSM (mutation cut off scanning matrix)(Pires et al., 2014b), DUET (Pires et al., 2014a), SDM (Worth et al., 2011) (Site Directed Mutator), SDM2 (Karnik et al., 2013), I-Mutant (Capriotti et al., 2005), MUpro (Parthiban et al., 2006), and the Cologne University Protein Stability Analysis Tool (CUPSAT)(Parthiban et al., 2006). These tools have different input requirements; mCSM, DUET, SDM, and CUPSAT require the protein’s three-dimensional structure, whereas MUpro simply needs the amino acid sequence. Additionally, I-Mutant was utilized to predict stability changes due to single-site mutations, with the tool capable of making predictions using either the protein sequence or structure based on SVM models.

### 2.11 Evolutionary Conservation Analysis

Evolutionary conservation analysis was performed using the ConSurf web server (Glaser et al., 2003). The Position-Specific Iterative Basic Local Alignment Search Tool (PSI-BLAST) was used to identify homologous sequences for the EDA, WNT10A, PAX9, and TSPEAR protein sequences, selecting the top 150 sequences for further analysis. Conservation scores of amino acid residues were calculated by ConSurf, which applies Bayesian inference to evaluate evolutionary conservation. The scores range on a nine-point scale, where a score of 1 represents low conservation, indicating variable positions, and a score of 9 indicates high conservation, representing conserved regions.

### 2.12 Structural Analysis, Modeling, and Validation

The 3D structures of PAX9, EDA, TSPEAR, and WNT10A proteins were obtained using AlphaFold(Jumper et al., 2021). Mutant models were generated through homology modeling based on AlphaFold templates (PAX9 AF-P55771-F1-v4, EDA AF-Q92838-F1-v4, TSPEAR AF-Q8WU66-F1-v4 and WNT10A AF-Q9GZT5-F1-v4) using the Swiss-Model webserver (Kiefer et al., 2009). To refine the models and minimize potential structural inconsistencies, energy minimization was performed using the “steepest descent” method with the GROMACS 2018 software suite (Berendsen et al., 1995). The quality and integrity of the modeled structures were further evaluated using the PDBsum tool (Laskowski et al., 2018).

### 2.13 RNA Sequence Retrieval

The RNA sequences under analysis were sourced from the following transcript IDs: *WNT10A* NM_025216, EDA NM_001399, and *TSPEAR* NM_144991. Short 41-nucleotide RNA segments were extracted with the mutation site precisely positioned at the 21st nucleotide having a sequence length of 20 bases on either side of the mutation site.

### 2.14 Prediction of RNA Secondary Structure

For predicting the secondary structures (2D) of the RNA snippets, the RNAStructure Web Server (version 6.0.1) was employed(Devi, Ranjan, Raj, et al., 2024)(Bellaousov et al., 2013). This server utilizes algorithms grounded in thermodynamic principles to forecast RNA secondary structures. These predictions elucidate folding patterns and potential base pairing interactions within the RNA molecules.

### 2.15 Assessment of Mutation Effects on RNA Structure

To explore the structural alterations induced by each missense mutation, the MutaRNA tool facilitated the mutational analysis of RNA sequence (Miladi et al., 2020). This analysis encompassed evaluating the intra-molecular base pairing potential, determining base pairing probabilities of the mutant RNA, and assessing accessibility (single-strandedness) in comparison to the wild-type counterparts (Miladi et al., 2020)(Bernhart et al., 2011). By incorporating remuRNA (Salari et al., 2013) and RNAsnp, this tool provides insights into the mutation-induced changes in RNA structures.

### 2.16 Prediction of Post-Translational Modifications (PTMs)

To identify potential PTMs in proteins, various computational tools were used. NetPhos 3.1(Blom et al., 1999) predicted phosphorylation sites (serine, threonine, tyrosine), while NetNGlyc (Pugalenthi et al., 2020) and NetOGlyc(Hansen et al., 1998) identified N- and O-glycosylation sites, respectively. GPSSNO(Xue et al., 2010) was employed for S-nitrosylation prediction, and GPSSUMO (Zhao et al., 2014) identified sumoylation sites and motifs. PrePS analyzed prenylation types, including farnesylation and geranylgeranylation (Maurer-Stroh & Eisenhaber, 2005). UbPred(Radivojac et al., 2010) predicted ubiquitination sites, Myristoylator identified N-terminal myristoylation, and CKSAAPPalm (Ranjan & Das, 2023) along with eqPalm were used for palmitoylation. Finally, GPS PAIL (Deng et al., 2016) predicted lysine acetylation sites. Each tool relied on protein sequence inputs to detect possible modifications, providing insights into how these PTMs might regulate protein function and stability, with implications for congenital tooth agenesis.

### 2.17 Molecular Simulations

GROMACS was used to perform molecular dynamics (MD) simulations in order to examine the structural behavior of the wild-type and mutant forms of the proteins PAX9, EDA, TSPEAR, and WNT10A. For these simulations, the GROMOS96 54a7 force field (Schmid et al., 2011)(Devi, Ranjan, & Das, 2024) was used. SPC216 water molecules were used to encircle the proteins in a dodecahedron box arrangement, with the proteins and box edges spaced at least 1.0 nm apart. In order to simulate physiological circumstances, 0.15 M NaCl and Na ions were added to electrically neutralize the system.

The steepest descent approach was used to minimize energy and resolve any steric conflicts. Subsequently, the system underwent two stages of equilibration: first, at 300 K, under the NVT ensemble (constant number of particles, volume, and temperature); and secondly, at 1 atm for 100 ps, under the NPT ensemble (constant number of particles, pressure to temperature). MD simulations were prolonged to a period of 50 ns following the equilibration phases. All mutant and wild-type structures achieved a stable equilibrium state within 10 ns. GROMACS tools (e.g., gmx rms, rmsf, gyrate, hbond, and sas) were used to analyze key parameters, such as solvent-accessible surface area (SAS), hydrogen bonds (intra-protein and protein-water interactions), radius of gyration (Rg), root mean square deviation (RMSD), and root mean square fluctuation (RMSF). The results were visualized using XMGRACE (Cowan & Grosdidier, 2000) and MS excel(Berk & Carey, 1998).

### 2.18 Principal Component Analysis (PCA) of Biophysical Metrics and Matplot analysis

To assess and visualize variability among different protein variants, we conducted Principal Component Analysis (PCA) based on key biophysical metrics: root mean square deviation (RMSD), root mean square fluctuation (RMSF), solvent-accessible surface area (SAS), hydrogen bond count (H-bonds), radius of gyration (Rg), and subcellular localization properties. These metrics were measured for a variety of wild-type and mutant proteins, including EDA (EDA_WT, EDA_R156H), PAX9 (PAX9_WT, PAX9_Q145*), TSPEAR (TSPEAR_WT, TSPEAR_419fs150, TSPEAR+Compound_Hetero), and WNT10A (WNT10A_WT, WNT10A_V145M, WNT10A_A135S).

To ensure fair comparison, the data were standardized, allowing each feature to contribute equally to the analysis and preventing any single metric from dominating the results. PCA was performed using the scikit-learn library(Kramer, 2016), reducing the dataset to two principal components (PC1 and PC2) that captured most of the variance.

The PCA results were visualized as a scatter plot, where each point represented a protein variant. Different colors and annotations were used to distinguish between the variants, facilitating easy identification of patterns and clusters. The protein variant analysis was conducted using Python (v3.12) (Python, 2021)) with pandas for data management and matplotlib (Hunter & Dale, 2007) for visualization. The biophysical metrics (RMSD, RMSF, SAS, H-Bonds, Rg) were plotted across the different protein variants using subplots. Distinct colors and triangular markers were employed for each metric, enhancing clarity and comparison.

### 2.19 Others relevant miscellaneous analysis

For the analysis of protein variants, several computational tools were employed. Secondary structure prediction and Gene Ontology (GO) annotations for wild-type and variant forms of PAX9, EDA, WNT10A, and TSPEAR were performed using the Psipred Workbench (Buchan & Jones, 2019). The protein sequences served as input for these predictions, providing insights into structural changes and functional implications associated with each variant. The difference distance matrix and Root Mean Square Deviation (RMSD) calculations were conducted using Superpose Version 1.0 (Bauer et al., 2008), enabling a detailed comparison of structural variations between the wild-type and mutant proteins. Additionally, TBtools (C. Chen et al., 2020), a versatile toolkit for biological data analysis, was utilized to generate heat maps, offering a clear visualization of the data across multiple metrics. Subcellular localization of the wild-type and variant proteins (PAX9, EDA, WNT10A, TSPEAR) was predicted using DeepLoc 2.0 (Thumuluri et al., 2022). Protein sequences were submitted to the DeepLoc 2.0 web server, which employs a convolutional neural network to predict localization probabilities for various cellular compartments (e.g., nucleus, cytoplasm, mitochondria). The predictions were analyzed to identify the most likely localizations for each variant, and results were visualized for comparative analysis.

Discover Studio(Systèmes, 2016) was used for detailed structural analysis, providing insights into the biophysical properties of each protein variant.

## 3 Result

### 3.1 WES Analysis

The WES analysis of CTA patients identified several variations in candidate genes linked to tooth agenesis. In ***WNT10A***, a homozygous variation V145M (c.433G>A) was found, classified as likely pathogenic with autosomal recessive (AR) inheritance, and is a known variant. Additionally, a heterozygous novel variant A135S (c.403G>T) was identified as a variant of uncertain significance (VUS) associated with autosomal dominant (AD) inheritance. For ***EDA***, a known homozygous variant R156H (c.467G>A), classified as likely pathogenic, was observed, linked to ectodermal dysplasia and X-linked recessive (XLR) inheritance. In ***PAX9*** a stop-gained mutation (c.433C>T; p.Gln145*), classified as pathogenic, known, and related to AR inheritance. In ***TSPEAR***, two compound heterozygous variations were identified: L219P (c.656T>C), a novel VUS with autosomal recessive (AR) inheritance, and a frameshift variant I419Lfs*150 (c.1255delA), classified as a known VUS. These findings provide insights into the genetic basis of tooth agenesis, highlighting both known pathogenic variants and novel uncertain variants for further investigation **(Table 1, Figure1).** Further Novel variants cross validated by sanger sequencing showed in Figure2.

**Table1:**
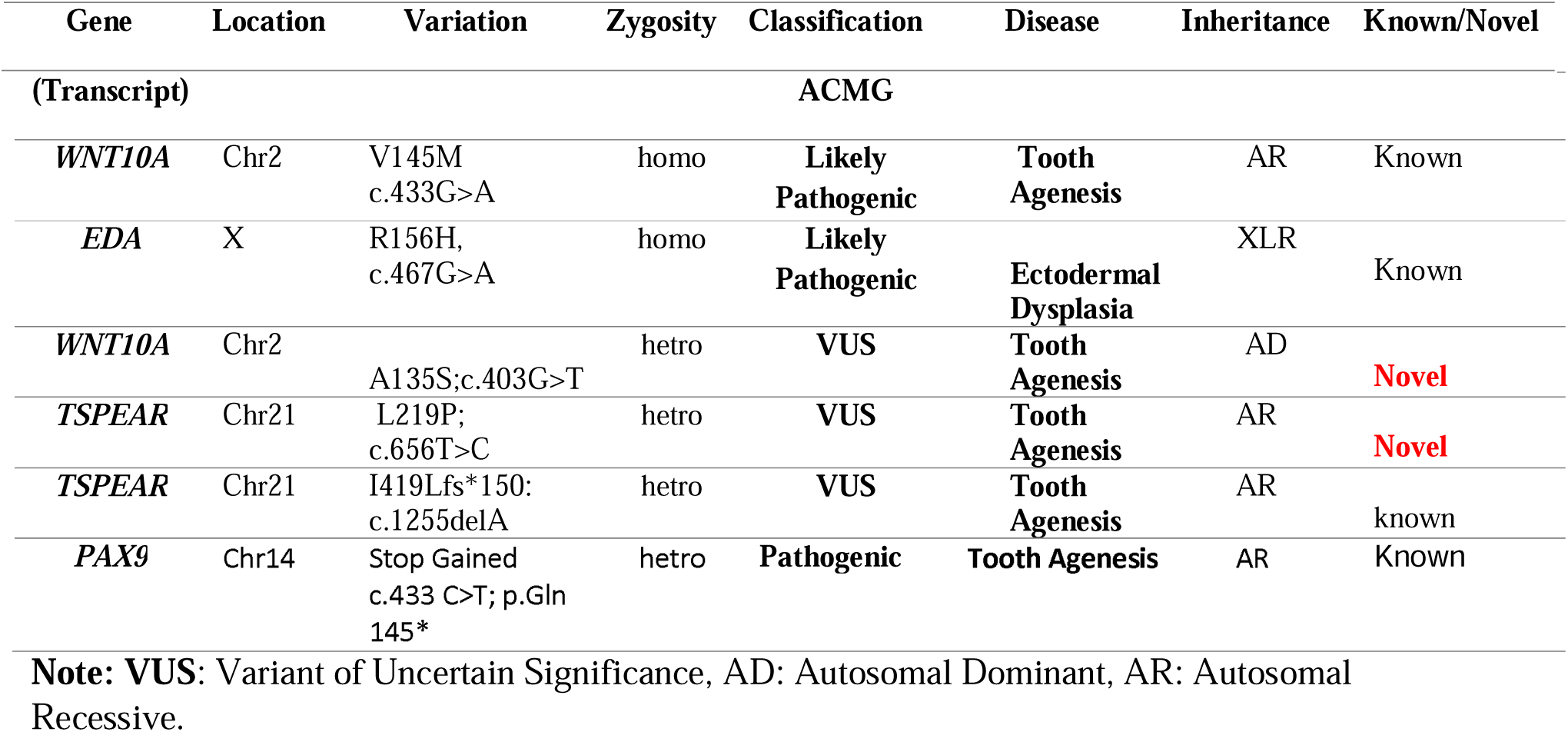
Screening of Known and Novel Variations in Candidate Genes in CTA Patients.

**Figure1:**
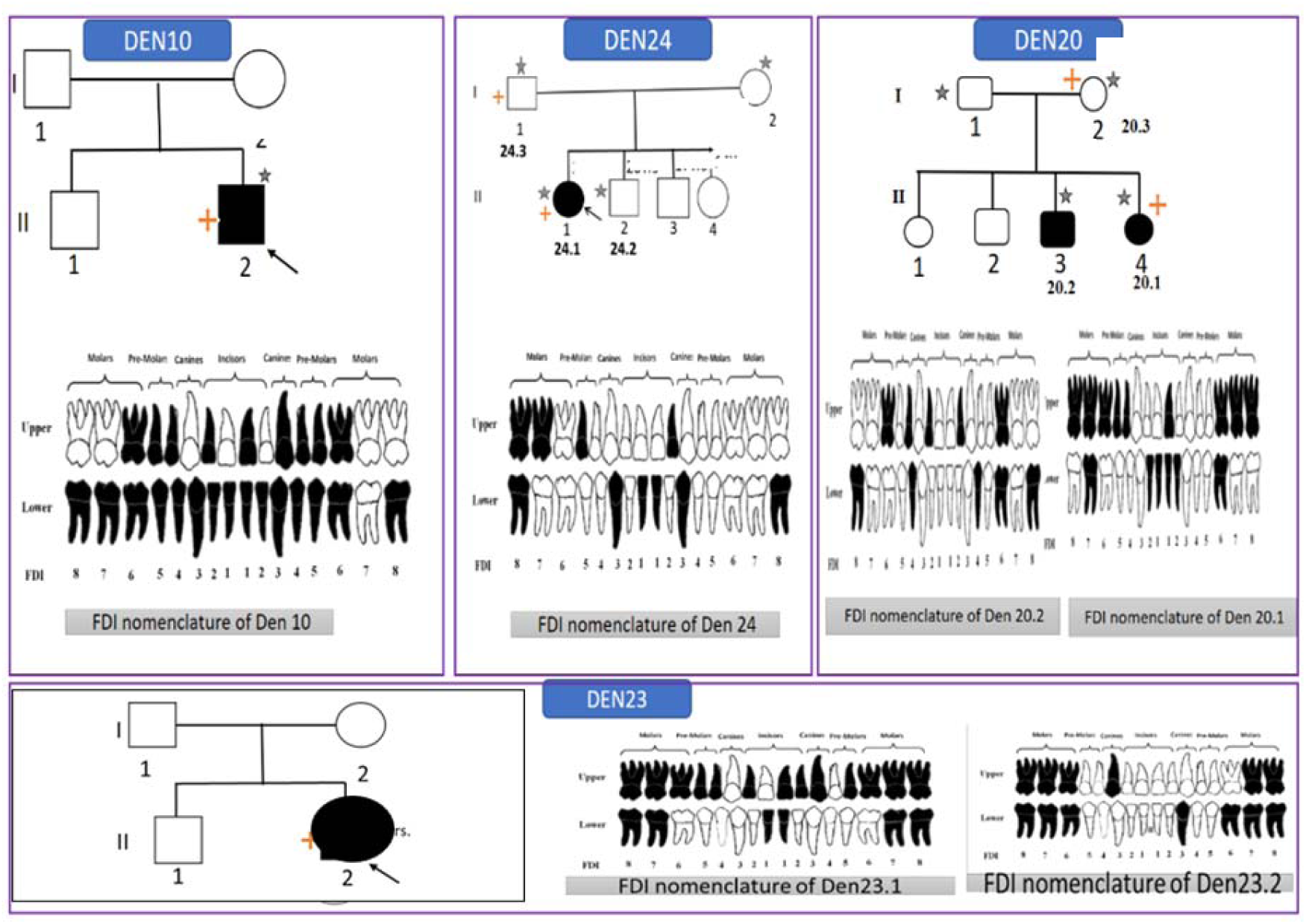
Clinical and Genetic Analysis of Patients with Tooth Agenesis. Panels (A-D) present clinical and genetic data from patients with tooth agenesis, showing key variants identified in *EDA, WNT10A, TSPEAR*, and *PAX9* genes: **(A) DEN10:** Pedigree with an *EDA* variant (p.I156H; c.467G>A) and OPG showing dental status. **(B) DEN24:** WNT10A variant (p.A135S; c.403G>T) with OPG and FDI dental mapping. **(C) DEN20:** TSPEAR variant (p.I219P; c.656T>C) in two family members, with corresponding OPGs. **(D) DEN23:** PAX9 variant (p.Gln45Ter; c.433C>T) with OPGs for two affected individuals. Each panel includes the patient’s pedigree, OPG, and FDI tooth chart to illustrate genotype-phenotype correlations. To maintain confidentiality and anonymity, each sample was assigned a unique study code. Readers are encouraged to contact the corresponding author for access to patients opg.

The variant analysis summarized here corresponds to the findings presented in **Table 2**. Each variant, including *TSPEAR* (L219P and I419fs*150), PAX9 (Q145\**), *WNT10A* (A135S and V145M), and *EDA* (R156H), has been assessed across multiple databases and predictive tools. The results indicate their potential pathogenicity, with entries in databases like OMIM, EXAC, and gnomAD, along with consistent damaging predictions from tools such as SIFT, Polyphen-2, MutationTaster2, and CADD3 Phred scores.

**Table2.** Analysis of Gene Variants Across Databases and Prediction Tools.

| Variants | Database reports |  |  |  |  |  | Predictions (scores) |  |  |  |
| --- | --- | --- | --- | --- | --- | --- | --- | --- | --- | --- |
|  | 1000<br>genome<br>s | EXA<br>C | gnomAD | Turkish<br>Variome | OMIM | ClinVar | SIFT | Polyphen-2 | Mutatio<br>nTaster<br>2 | CADD3<br>Phred<br>score |
| *TSPEAR*:<br>L219P;c.656T>C | NA | NA | NA | NA | 603140 | NA | NA | NA | NA | 26 |
| *TSPEAR*:<br>I419fs*150<br>:<br>c.1255delA | NA | <0.01 | <0.01 | NA | 603140 | NA | NA | NA | D | NA |
| *PAX9*:<br>Q145*;c.433C>T | NA | NA | NA | NA | 128400 | NA | D | D | D | D |
| *WNT10A*<br>:A135S;<br>c.403G>T | NA | NA | NA | NA | 608522 | NA | D | D | D | 24.4 |
| *WNT10A*<br>(V145M)<br>c.433G>A | NA | 0.0025 | 0.0073 | 0.0446% | 606268 | Likely<br>Pathogenic | D | D | D | 25.1 |
| *EDA*:<br>p.R156H;<br>c.467G>A | NA | NA | 0.0009 | 0.0356 | 129920 | NA | D | D | D | 26.8 |

**Notes:** This table summarizes variant details, including relevant database reports and pathogenicity predictions. "NA" signifies data unavailability. The prediction scores indicate potential impacts on protein function: "D" denotes a damaging prediction by SIFT, PolyPhen-2, and MutationTaster2, while the CADD3 Phred score highlights variants with higher pathogenic potential, generally above 20.

### 3.2 RT-PCR Assessment of Novel Variants Affecting Wnt and EDA Signaling

RT-PCR analysis revealed significant reductions in the expression levels of *EDA* and *WNT10A* in samples carrying the identified variants. Specifically, *EDA* expression was notably decreased compared to the internal control (Figure 2. G), with ΔCT values indicating substantial downregulation. Similarly, *WNT10A* expression was significantly lower in variant samples (Figure 2. H), further supported by the ΔCT values. Statistical analysis confirmed these findings, with ****p < 0.0001 for *EDA* and ***p < 0.001 for *WNT10A*, indicating that the identified variants adversely affect EDA and WNT signaling pathways, potentially contributing to the observed phenotypes.

**Figure2:**
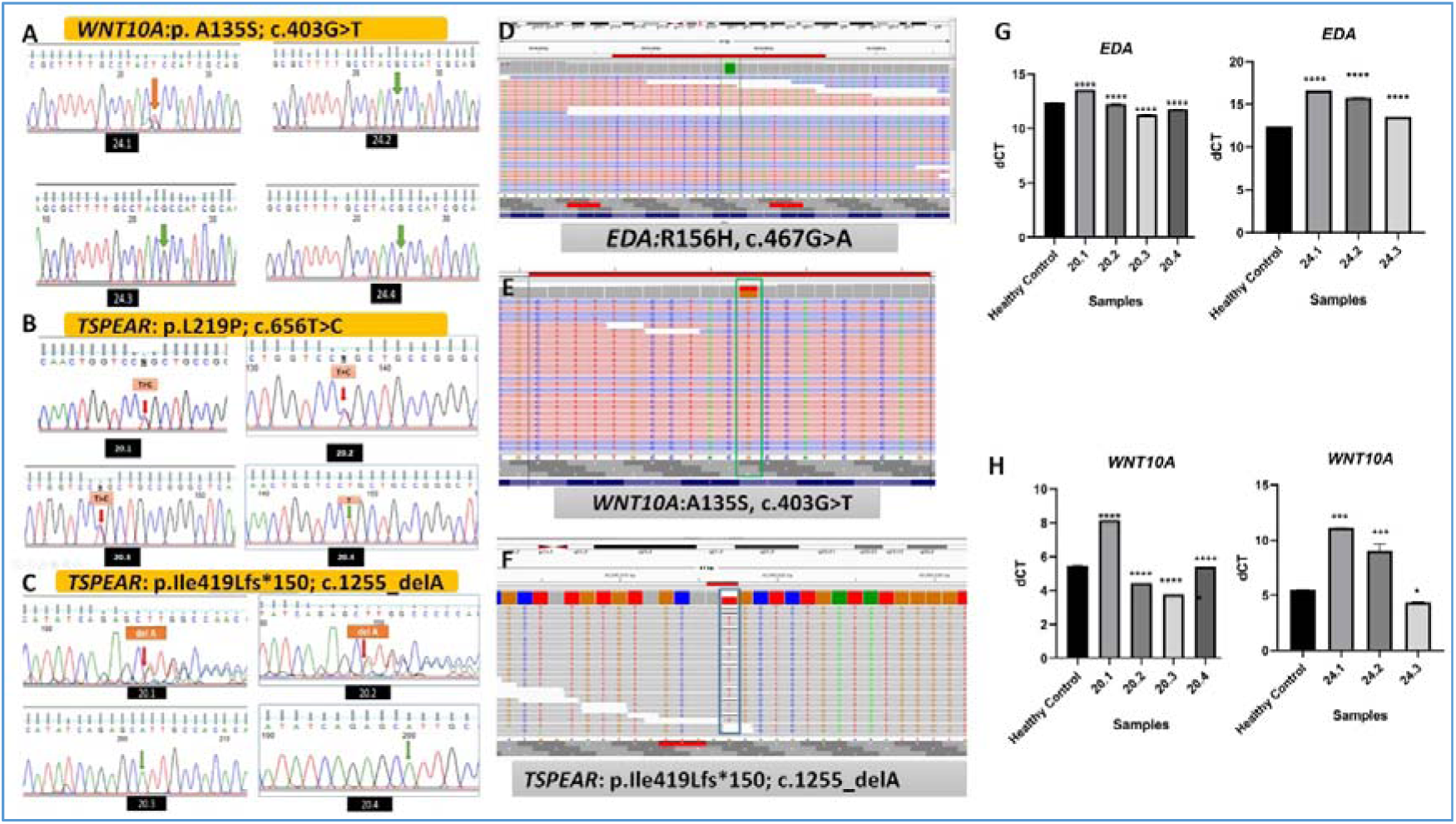
Validation of genetic variants and their impact on *EDA* and *WNT* signaling through gene expression. (A–C) Sanger sequencing chromatograms showing validated variants in the genes *WNT10A* (p.A135S; c.403G>T) (A), *TSPEAR* (p.L219P; c.656T>C) (B), and *TSPEAR* (p.Ile419fs*150; c.1255_delA) (C). The red arrows indicate the locations of nucleotide changes, while green arrows show nucleotides that remain unchanged. (D–F) Integrative Genomics Viewer (IGV) screenshots showing the presence of these variants in the sequencing data. Panel (D) shows the *EDA* variant (R156H; c.467G>A), (E) shows the *WNT10A* variant (A135S; c.403G>T), and (F) shows the *TSPEAR* frameshift mutation (p.Ile419fs*150; c.1255_delA). (G–H) RT-PCR analysis of *EDA* and *WNT10A* gene expression levels. (G) Relative expression levels of *EDA* in samples carrying the variant compared to the internal control, showing a significant reduction in expression. (H) Relative expression levels of *WNT10A* in samples with the variant, also showing a significant decrease in expression. Δ**CT** values are shown for each sample, with statistical significance indicated as follows: ****p < 0.0001, ***p < 0.001. These findings indicate that the identified variants affect *EDA* and *WNT* signaling pathways, potentially contributing to the observed phenotypes.

### 3.3 WES Gene Panel

Data from five control samples and six CTA patients were filtered using a minor allele frequency (MAF) criterion of ≤20% as part of the full WES analysis. The emphasis was on possible related genes unique to the CTA patients by excluding common variations using those found in the control samples. 709 genes made up the final curated gene panel that was discovered using this method. Of the genes in the panel, 21 are consistently seen in all six patients, 391 are shared by three patients, 204 are found in four patients, and 98 are seen in five patients. By giving priority to genes found in more than three CTA samples, the gene panel was improved in terms of specificity and CTA relevance.

Gene panel showed in Supplementary File Information S2. Ten genes (*TM4SF20, LCORL, OR4F21, TNS2, VWA8, MYH6, KNL1, LOC400499, MRTFB,* and *PRR32*) contained mutations categorized as synonymous, non-synonymous, or found in UTR sections, according to a thorough study of variants present in 21 genes shared by all six patient samples. This analysis did not include variants that fell outside of these categories (Table 3). Diverse conclusions were drawn from the examination of variations in ten genes that were present in six patient samples. Variants in *MYH6* (G56R) and *TM4SF20* (A27V) were found to be benign, had moderate minor allele frequencies (MAF), and were present in several samples, indicating that they were common.

**Table 3:** Analysis of genes identified across all six tooth agenesis patients, highlighting the presence of different variants within each gene.

| Gene | Function/Diseases | Variant/AA Position | ACMG | MAF/NOVEL | HGVSC | Effect | Samples Present |
| --- | --- | --- | --- | --- | --- | --- | --- |
| **TM4SF20** | Tetraspanin, cell adhesion/**Autism, AZ** | A27V | Benign | 10.68% | c.80C>T | NON_SYNONYMOUS_CODING | DEN1, DEN10, DEN20, DEN12 |
| **LCORL** | Transcriptional regulator/**Growth Dev., AZ** | L1734P | VUS | **Novel** | c.5201T>C | NON_SYNONYMOUS_CODING | DEN1 |
| **LCORL** | Transcriptional regulator/**Growth Dev., AZ** | P1709R | VUS | <0.01% | c.5126C>G | NON_SYNONYMOUS_CODING | DEN1 |
| **LCORL** | Transcriptional regulator/**Growth Dev., AZ** | T1042 | Benign | 13.14% | c.3126T>C | SYNONYMOUS_CODING | DEN10, DEN20.1, DEN24.1, DEN12 |
| **OR4F21** | Olfactory receptor/**No diseases reported** | K310R | VUS | **Novel** | c.929A>G | NON_SYNONYMOUS_CODING | DEN1, DEN10, DEN20.1, DEN24.1, DEN12, DEN23.1 |
| **OR4F21** | Olfactory receptor/**No diseases reported** | F44L | VUS | **Novel** | c.130T>C | NON_SYNONYMOUS_CODING | DEN1, DEN20.1, DEN12 |
| **TNS2** | Tensin protein, cell migration/**Kidney Diseases** | I858M | VUS | 0.02% | c.2574C>G | NON_SYNONYMOUS_CODING | DEN1 |
| **TNS2** | Tensin protein, cell migration/**Kidney Diseases** | G774E | Benign | 0.11% | c.2321G>A | NON_SYNONYMOUS_CODING | DEN12 |
| **VWA8** | Von Willebrand factor type A domain/**Covid19, Cleft Lip Palate, Autism** | R552H | Benign | 5.53% | c.1655G>A | NON_SYNONYMOUS_CODING | DEN1 |
| **VWA8** | Von Willebrand factor type A domain/**Covid19, Cleft Lip Palate, Autism** | I1439L | Benign | 1.44% | c.4315A>C | NON_SYNONYMOUS_CODING | DEN10 |
| **MYH6** | Myosin heavy chain, cardiac muscle/**CHD** | G56R | Benign | 7.83% | c.166G>A | NON_SYNONYMOUS_CODING | DEN10, DEN12 |
| **KNL1** | Kinetochore protein/**Mic rocephaly, Cancer** | A1270V | VUS | 0.01% | c.3809C>T | NON_SYNONYMOUS_CODING | DEN20.1 |
| **LOC400499** | Hypothetical protein/**Unknown** | G2806R | NA | NA | c.8416G>A | NON_SYNONYMOUS_CODING | DEN1 |
| **LOC400499** | Hypothetical protein/**Unknown** | I176V | NA | NA | c.526A>G | NON_SYNONYMOUS_CODING | DEN20.1 |
| **MRTFB** | Myocardin-related transcription factor/**Autism, CHD** | A135 | Benign | 7.88% | c.405T>C | SYNONYMOUS_CODING | DEN1, DEN10, DEN20.1, DEN24.1, DEN12, DEN23.1 |
| **PRR32** | Proline-rich protein/**Intellectual Development Disorder** | M193T | Benign | 9.89% | c.578T>C | NON_SYNONYMOUS_CODING | DEN20.1, DEN24.1 |

**Note:** This table presents gene variants, their functions, and disease associations across samples. "VUS" (variant of uncertain significance) requires further study for clinical impact. "NOVEL"
denotes newly observed variants. "Benign" and "NA" Not Available. Highlighted showed present in all samples. To maintain confidentiality and anonymity, each sample was assigned a unique study code.

*LCORL* revealed three variations, including a benign synonymous variant (T1042) that was found in several samples and two variants of unclear significance (VUS) L1734P (new) and P1709R (rare), suggesting possible variation in regulatory functions. Two novel VUS (K310R, F44L) were found in *OR4F21* across samples, suggesting unique variants of unknown importance. There were two non-synonymous variations found in *TNS2* (I858M and G774E), the former of which was categorized as VUS and the latter as benign. With modest MAFs, the *VWA8* (R552H, I1439L) and *PRR32* (M193T) variants were likewise innocuous.

Although there was no clinical categorization information available, *LOC400499* had two non-synonymous variations (G2806R, I176V), and *KNL1* displayed a rare VUS (A1270V). A harmless synonymous variation (A135) of *MRTFB* was found in several samples, indicating a shared polymorphism. All things considered, the study found both new and benign VUS variations, offering information on the genetic causes of a number of illnesses. *TM4SF20* (autism, Alzheimer’s), *LCORL* (growth disorders, Alzheimer’s), *OR4F21* (no known diseases), *TNS2* (kidney diseases), *VWA8* (COVID-19, cleft lip palate, autism), *MYH6* (congenital heart disease (CHD)), *KNL1* (microcephaly), *LOC400499* (unknown), *MRTFB* (autism, CHD), and *PRR32* (intellectual development disorders) were among the gene-disease associations found by the analysis.

These correlations point to possible genetic causes of the disorders seen in the patient samples. In all six samples, two variations were consistently found: *MRTFB* (A135, c.405T>C) and *OR4F21* (K310R, c.929A>G). Whereas the *MRTFB* variation is a known synonymous alteration, the *OR4F21* variant is a new, non-synonymous modification.

Key pathways and functions were discovered by the WES panel’s enrichment analysis (Figure 3). Pathway study revealed important roles in apoptotic signaling, cell cycle control, and DNA damage response. Protein localization to membranes and DNA replication and repair were examples of biological activities that suggested roles in cellular growth and stability. Mitochondrial complexes, CMG complexes, and cell junctions were found to be abundant in cellular components, indicating their significance in energy metabolism, DNA synthesis, and cell communication. DNA binding, helicase activity, and oxidoreductase activity were the main molecular activities that were linked to enzymatic processes and genomic stability. Human phenotypic ontology revealed links to diseases such as myelodysplasia and bone marrow hypoplasia, suggesting genetic susceptibilities to hematological and developmental disorders.

**Figure3:**
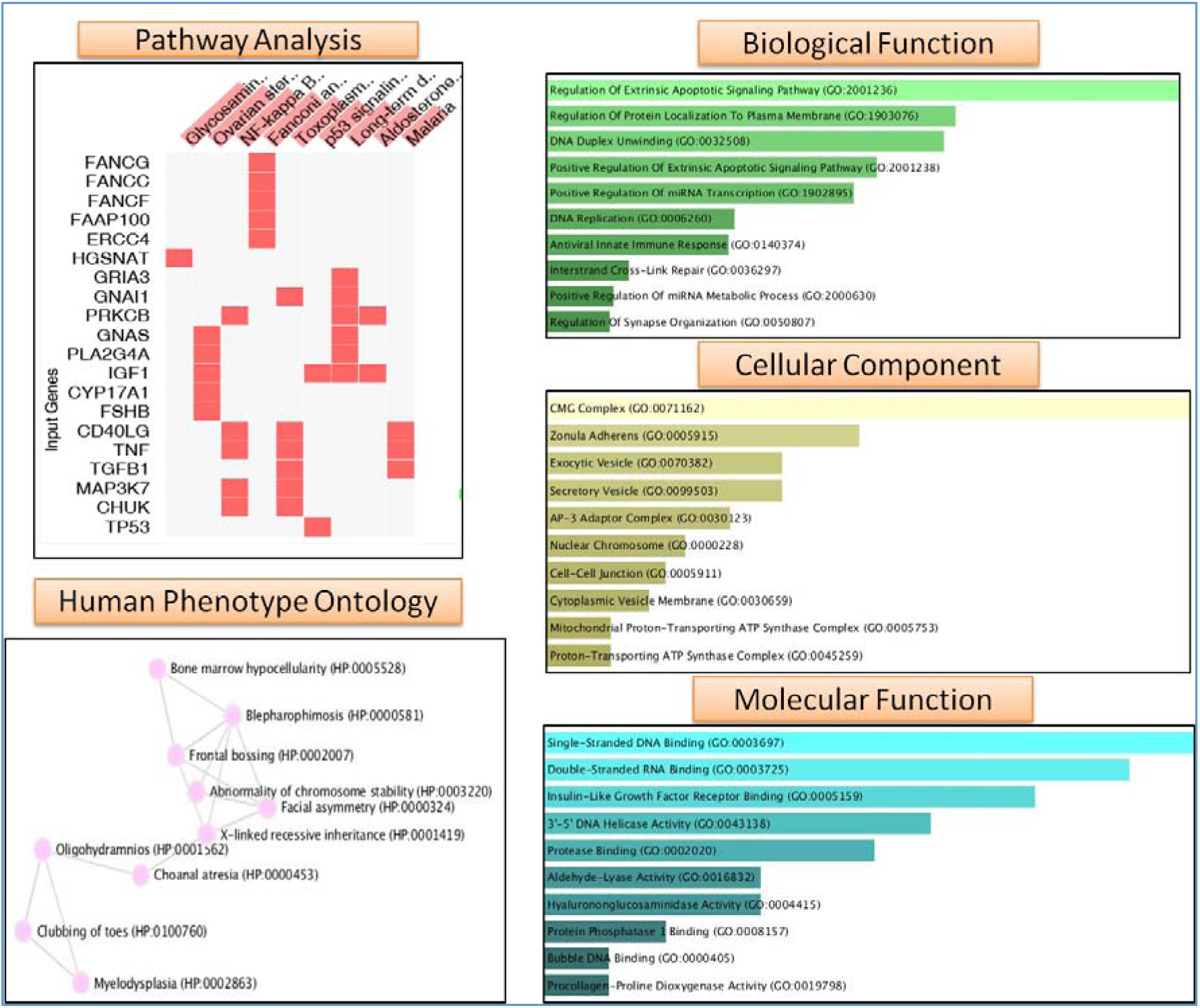
Enrichment analysis of the WES gene panel. The figure displays the results of enrichment analysis for the identified genes in terms of biological processes, cellular components, and molecular functions. The bars represent the enrichment score for each GO term, indicating the significance of the gene’s involvement in the corresponding biological process, cellular component, or molecular function. The results highlight the potential roles of these genes in various cellular pathways and processes.

The PPI network analysis of the WES gene panel identified 470 nodes and 1211 interactions, showing a well-connected network structure **(Figure 4 Panel A).** The analysis revealed 11 network modules with key hub genes, including *TNF, TP53, STAT3*, and *CXCL12* (**Figure 4** Panel C), indicating their central role. Yellow-highlighted genes within the network represent those associated with developmental processes, suggesting their relevance in tooth development. Summary metrics (**Figure 4** Panel B) showed an average of 5.153 neighbors per node, a clustering coefficient of 0.126, and a network density of 0.005.

**Figure 4:**
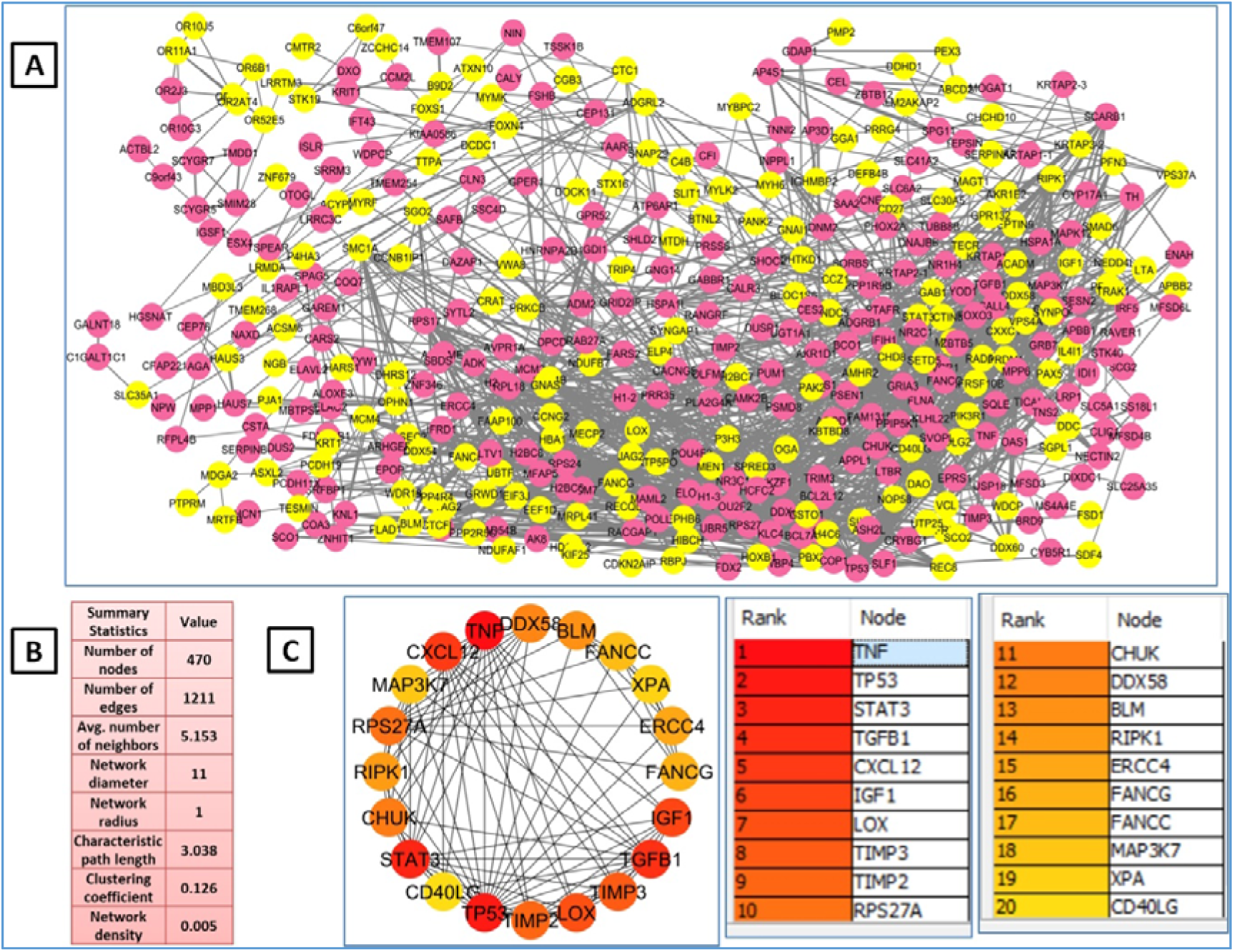
PPI network analysis of the WES gene panel. The figure represents the identified genes as nodes connected by edges, indicating protein-protein interactions. Genes involved in developmental processes are highlighted in yellow. The size of each node corresponds to its degree (number of connections), and the color represents its clustering coefficient. The top 20 hub genes (genes with the highest number of connections) are listed in a separate table. **Panel A:** The network visualization, where nodes represent genes and edges represent protein-protein interactions. **Panel B:** Summary statistics of the network, including the number of nodes, edges, average degree, diameter, radius, characteristic path length, clustering coefficient, and network density. **Panel C:** A circular representation of the top 20 hub genes, with their ranks and node names listed. The color gradient indicates the degree of each hub gene.

### 3.4 Multiomics Integration

The multi-omics integration analysis revealed shared gene elements across the WES Panel, GSE56486_UP, and GSE56486_Down datasets. The Venn diagram showed 676 unique genes in the WES Panel, 726 in GSE56486_UP, and 60 in GSE56486_Down, with 18 genes common between the WES Panel and GSE56486_UP, and 15 shared between the WES Panel and GSE56486_Down. The heatmap of the 18 common genes suggests possible functional or regulatory relationships. KEGG pathway analysis indicated that upregulated genes were enriched in pathways like NOD-like receptor signaling, steroid biosynthesis, and cytoskeletal metabolism, while downregulated genes were linked to pathways involving endocytosis, bacterial invasion of epithelial cells, and fatty acid degradation, hinting at disruptions in cellular metabolic activities **(Figure5).**

**Figure5:**
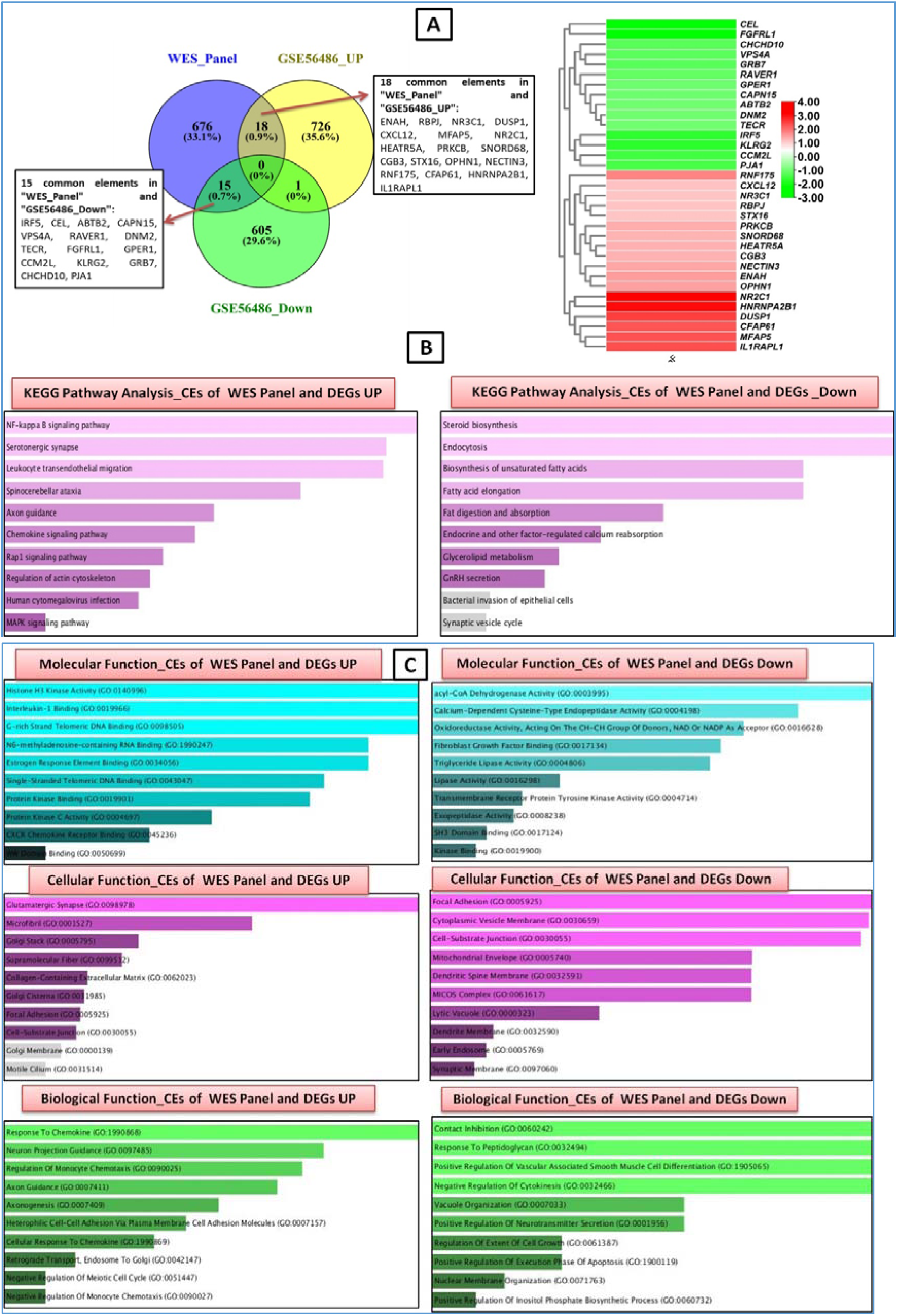
Venn Diagram, Heatmap, Pathway, and GO Enrichment Analysis of Common Elements Between WES Gene Panel and Differentially Expressed Genes (DEGs). **Panel A:** A Venn diagram illustrating the intersection between the WES gene panel and DEGs that are upregulated (GSE56486 UP) or downregulated (GSE56486 Down) within the dataset. The numbers in each segment denote the count of genes in each category. Accompanying this, a heatmap visualizes the common elements (CEs) shared between the WES gene panel and DEGs. Color intensity reflects the fold change, with shades of red indicating upregulation and shades of green indicating downregulation. **Panel B:** Pathway analysis of the DEGs, showcasing pathways enriched among the upregulated and downregulated genes. This highlights the pathways significantly affected by gene expression changes within the dataset. **Panel C:** Enrichment analysis of the common elements (CEs) between the WES gene panel and DEGs, showing results for three main Gene Ontology (GO) categories: Biological Processes (BP), Molecular Functions (MF), and Cellular Components (CC). Bars represent the enrichment score for each GO term, indicating the significance of the CEs in the relevant biological processes, molecular functions, or cellular components. This panel underscores the functional distinctions between upregulated and downregulated DEGs within the WES gene panel.

The functional analysis integrating WES panel data with differentially expressed genes (DEGs) shows that upregulated genes are primarily associated with binding activities and enzymatic functions, while downregulated genes are linked to oxidoreductase activities, cytoskeletal binding, and ion transport. On the cellular level, upregulated genes contribute to extracellular matrix components, ion transport, and adhesion, whereas downregulated genes are connected to cell junctions, protein complexes, and cytoskeleton components. In terms of biological function, upregulated genes are enriched in processes like cell adhesion, signal transduction, and vascular development, while downregulated genes are focused on cell proliferation, immune response, and metabolism **(Figure5).** A comprehensive analysis of 33 differentially expressed genes (DEGs) revealed several notable variants. Among these, *ENAH* exhibited a benign deletion variant (ERQERLD217D) in samples DEN1 and DEN20.1. *LOX* showed a benign non-synonymous variant (R158Q) with a minor allele frequency (MAF) of 17.02%. *NR3C1* and *TIMP2* presented synonymous changes, while *IRF5*, *CEL* (novel), and *ABTB2* had variants classified as variants of uncertain significance (VUS). Additionally, *RNF175*, *TGFB1*, and *PRKCB* presented benign and VUS variants across multiple samples, contributing to the understanding of genetic factors associated with various diseases. Further insights can be gleaned from the detailed **Table4.**

**Table 4:** Comprehensive Analysis of Variants in 33 DEGs Common to the WES Panel and Top 10 Hub Genes.

| Gene | Function/Diseases | AA Change | ACMG | MAF/NOVEL | CTA_DEGs | HGVSC | Effect | Samples |
| --- | --- | --- | --- | --- | --- | --- | --- | --- |
| ***ENAH*** | Actin polymerization regulator involved in cell motility and morphology/**Cancer, Nephropathy.** | ERQER LD217D | Benign | 9.59% | UP regulated | c.651_668 delGCGG CAGGAA CGCCTG GA | CODON_CHANGE_PLUS_CODON_DELETION | DEN1, DEN20.1 |
| ***LOX*** | Lysyl oxidase; catalyzes the cross-linking of collagen and elastin, crucial for extracellular matrix stability/**Cancer, Diabetic, Heart, ALS** | R158Q | Benign | 17.02% | NA (Top 10 hub gene) | c.473G>A | NON_SYNONYMOUS_CODING | DEN1, DEN10, DEN20.1 |
| ***NR3C1*** | Glucocorticoid receptor; regulates genes controlling development, metabolism, and immune response/**Diabetics, Nephrotic, Crohn Diseases, Heart, Schizophrenia.** | N766 | Benign | 15.02% | UP regulated | c.2298T>C | SYNONYMOUS_CODING | DEN1 |
| ***IRF5*** | Interferon regulatory factor; involved in innate immune response and regulation of inflammatory cytokines/**Auto immune, Bowel, Cancer, Kidney Diseases.** | I103V | VUS | <0.01% | Down regulated | c.307A>G | NON_SYNONYMOUS_CODING | DEN1 |
| ***CEL*** | Carboxyl ester lipase; important in the digestion and absorption of dietary fats./**Diabetic, Cancer, Kidney** | S654A | VUS | **Novel** | Down regulated | c.1960T>G | NON_SYNONYMOUS_CODING | DEN1, DEN20.1 |
| ***ABTB2*** | Ankyrin repeat and BTB (POZ) domain-containing protein; associated with cellular signaling pathways/**Cancer.** | H250Q | VUS | 7.92% | Down regulated | c.750C>G | NON_SYNONYMOUS_CODING | DEN1 |
| ***IGF1*** | Insulin-like growth factor 1; plays a key role in growth and development, especially in muscle and bone/**Cancer, Schizophrenia, Diabetes, Miscarriage.** |  | Benign | 9% | NA (Top 10 hub gene) | c.*6433G>C | UTR_3_PRIME | DEN1, DEN10 |
| ***TIMP2*** | Tissue inhibitor of metalloproteinase 2; inhibits matrix metalloproteinases (MMPs), which degrade the extracellular matrix/**Cancer.** | S101 | Benign | 12.54% | NA (Top 10 hub gene) | c.303G>A | SYNONYMOUS_CODING | DEN1, DEN10, DEN20.1 |
| ***RNF175*** | E3 ubiquitin-protein ligase; involved in protein degradation through ubiquitination/**Cancer.** | L307F | Benign | 18.5% | UP regulated | c.921G>C | NON_SYNONYMOUS_CODING | DEN20.1, DEN24.1, DEN12, DEN23.1 |
| ***TGFB1*** | Transforming growth factor-beta 1; regulates cell proliferation, differentiation, and development, as well as immune responses/**Kidney, Osteosclerosis, Heart, Cancer.** | R25P | Benign | 7.11% | NA (Top 10 hub gene) | c.74G>C | NON_SYNONYMOUS_CODING | DEN12 |
| ***PRKCB*** | Protein kinase C beta; plays a role in various cellular processes including proliferation, differentiation, and apoptosis/**Diabetes, Cancer.** | V139 | VUS | <0.01% | UP regulated | c.417G>A | SYNONYMOUS_CODING | DEN10 |
| **STAT3** | Signal transducer and activator of transcription 3; involved in cell growth and apoptosis regulation, playing a key role in cytokine signalling/**Inflammatory, Autoimmune Diseases.** |  | Benign | 7.5% | NA (Top 10 hub gene) | c.*893dup A | UTR_3_PRIME | DEN12 |
| **CXCL12** | Chemokine (C-X-C motif) ligand 12; mediates chemotaxis, especially involved in immune cell migration and development/**Cancer, Diabetes.** |  | NA | NA | UP regulated | c.*519G> A | UTR_3_PRIME | DEN1, DEN10, DEN12 |

**Note:** This table includes gene variants, functions, disease associations, and regulatory status across samples. "Benign" indicates non-pathogenicity, while "VUS" (variant of uncertain significance) may require further research. To maintain confidentiality and anonymity, each sample was assigned a unique study code.

### 3.5 Diseases Association

The genetic variants identified in patients with tooth agenesis show associations with several systemic diseases. *TM4SF20*, involved in cell adhesion, has been linked to autism and AZ. *LCORL*, a transcriptional regulator, is associated with growth development disorders, AZ, and autism. *OR4F21*, an olfactory receptor gene, presents novel variants but has no reported disease associations. *TNS2*, associated with kidney diseases, harbors mutations that could impact cell migration, possibly contributing to nephropathy and other organ-related conditions. *VWA8*, involved in extracellular matrix stability, is associated with COVID-19, cleft lip palate, and autism. *MYH6*, related to cardiac muscle function, is linked to CHD. The role of cellular division and mitosis in tooth and brain development is supported by *KNL1*, which is linked to microcephaly and cancer. A putative protein called LOC400499 may have important but unidentified roles in systemic illnesses. Autism and congenital heart disease are linked to *MRTFB*, which is involved in transcription and cell signaling. Lastly, there is more evidence linking neurological issues to tooth growth through *PRR32*, which is linked to intellectual development difficulties.

The integrity of the extracellular matrix depends on the cross-linking of collagen and elastin, which is catalyzed by the *LOX* gene (lysyl oxidase). It has a connection to ALS. The *LOX* R158Q variation, which occurs 17.02% of the time, is categorized as benign. This variation, which was shown to be among the top 10 hub genes, results in a non-synonymous coding mutation due to a nucleotide change at c.473G>A. Three patient samples include *LOX*.

### 3.6 Relative Entropy

The relative entropy scores (H(wt:mu)) computed by remuRNA in mutaRNA analysis for each coding sequence change in the investigated genes provides the information of structural impact of single nucleotide change within RNA sequences. These are summarized in Supplimentary Information S1. The higher the relative entropy value, the stronger the structural impact of the nucleotide-change on the RNA structure. The variant *WNT10A* c.403G>T stands out with a higher relative entropy score of 3.409, suggesting a substantial alteration in RNA structure compared to other variants examined followed by *EDA c.467G>A* variant and *TSPEAR c.656T>C* exhibiting relatively high entropy scores of 2.248 and 1.392, respectively, indicating notable structural impacts. In contrast, the *WNT10A c.433G>A* variant reports a lower relative entropy score of 0.355, suggesting a comparatively lesser impact on RNA structure.

### 3.7 Analysis of RNA structural features

Further analysis of the variants’ impact on RNA structural features revealed distinct impacts of single nucleotide polymorphisms (SNPs) on the wild type (WT) and mutant (MT) RNA sequences. The results indicate that the variant *WNT10A c.403G>T* has the most pronounced effect on RNA structure, followed by *TSPEAR c.656T>C*, whereas minor alterations are observed in *WNT10A c.433G>A* and *EDA c.467G>A* (Figure 6, 7). Similarly, these alterations in the base pairing probabilities and change in structure can be observed in 2D structures of WT and MT depicted in Supplementary Information S1. In Figure 6, heat map dot plots illustrate the base pairing potential of both WT and MT RNAs, with darker color dots indicating higher probabilities of base pairing. Circular plots depict the same base pair probabilities, with darker hues of grey indicating higher probabilities. Analysis of changes in base pair probabilities, as shown in Figure 6 (Pr (bp in WT) – Pr (bp in mut)), highlights difference in base pairing patterns between WT and MT RNAs at specific locations. The weakened base pairing potential and increased probabilities of base pairing within each MT RNA are depicted in Figure 7.

**Figure 6:**
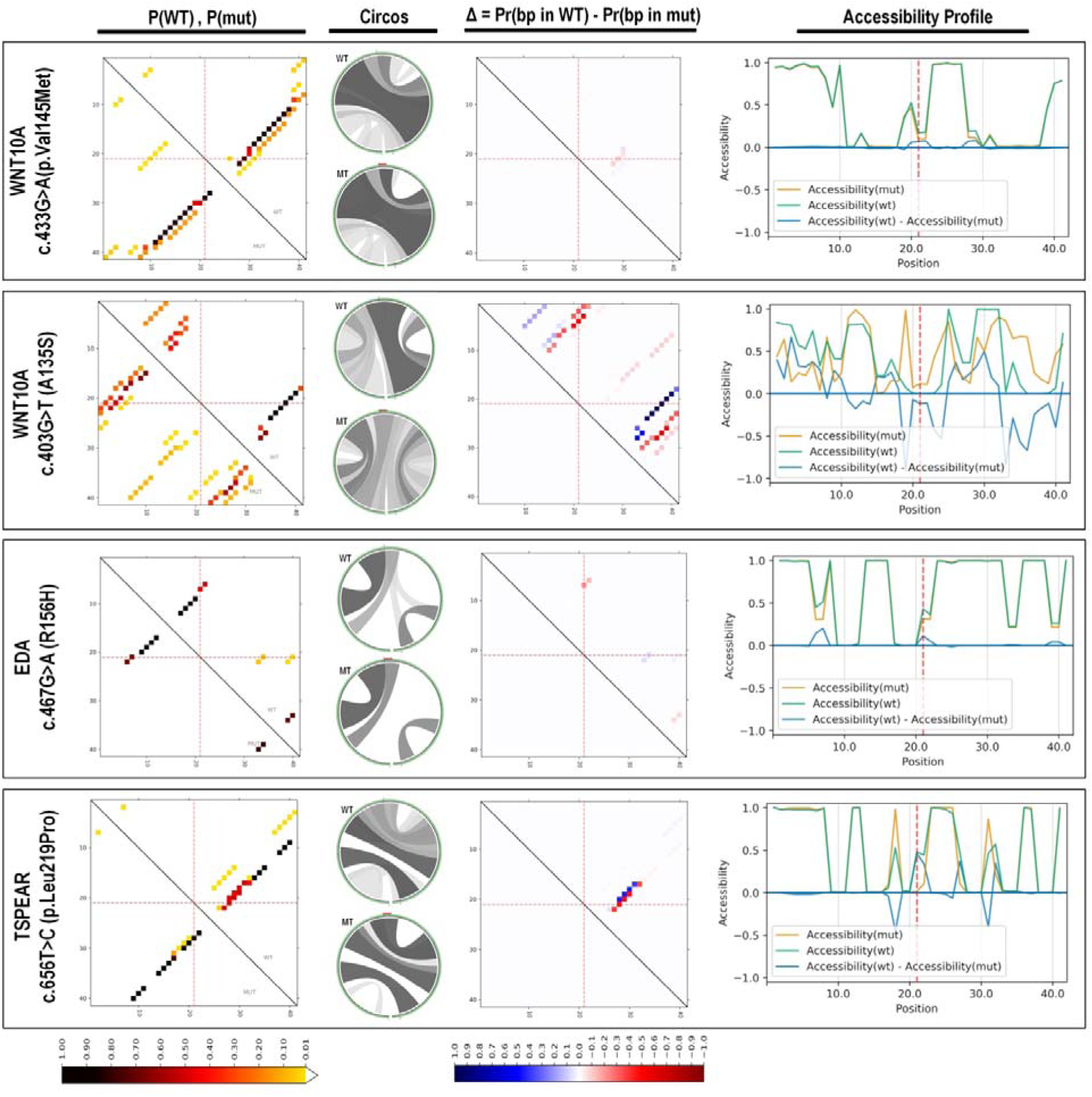
Analysis of RNA Structural Features. This figure presents analysis of RNA structural features for both wild type (WT) and mutant (MT) RNA sequences of each variant using mutaRNA tool. The mutated position is indicated by red dotted lines or mark. The first heat map like dot plots with the top-right and bottom-left panels provides a heatmap-like representation of base pairing probabilities in wild-type (p(WT)) and mutant (p(MT)) RNA sequences. The darker dots indicate higher probabilities of base pairing. The upper and lower circos plots visualize the base pair probabilities (p(WT) and p(MT)), with the sequence beginning from the 5′ end positioned at the bottom slight-left and extending clockwise until reaching the 3′ end. The variant of interest, located at position 21, is indicated by a red mark positioned on the top of the mutant circular plot. Higher probabilities are represented by darker gray scales. The dot plot on the right of circos shows the differences in base pairing probabilities between the mutated and wild-type RNA Pr (bp in WT) – Pr (bp in mut)). In this, base pairs weakened by the mutation are shaded in blue, while regions exhibiting increased base pair probabilities in the mutant are highlighted in red. The accessibility profiles of WT and MT RNA sequences, in terms of unpaired probabilities, are depicted on the right. The blue line illustrates the variations in accessibility (WT-mut), where negative values indicate positions more prone to being unpaired in the mutant compared to the wild type (WT).

**Figure 7:**
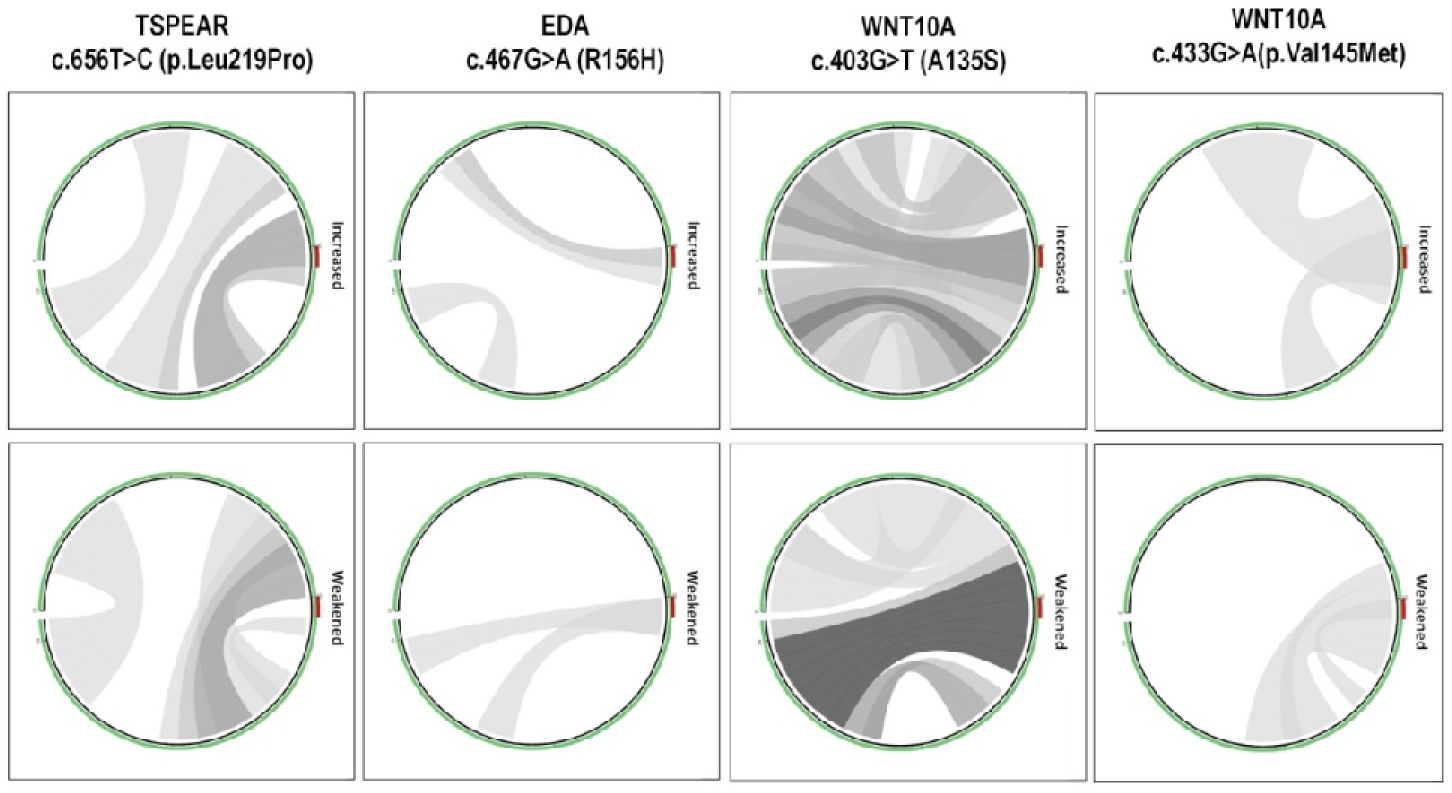
Comparative Visualization of Mutation Effects. The comparative circos plots in this figure aid in the easy identification and comparison of mutation effects. Right and left circus represent the increased base pairs and weakened base pairs probabilities between the nucleotides within the RNA sequence. Darker shades of gray indicate higher absolute changes. The variant position is marked by a red bar in the plot.

The accessibility profiles, as depicted in Figure 6, reveal varying effects of the examined variants on RNA. The accessibility profiles are assessed based on RNA single-strandedness (unpaired probabilities), representing the likelihood of each nucleotide position being unpaired within the RNA sequences. It is a crucial factor influencing RNA-protein or RNA-RNA interactions. Notably, the variant *WNT10A c.403G>T* exhibits the most pronounced alteration in accessibility, following closely, the variant *TSPEAR c.656T>C* also shows notable changes in accessibility profiles, indicating an impact on RNA single-strandedness. In contrast, minor alterations in accessibility profiles are observed for the variants *WNT10A c.433G>A* and *EDA c.467G>A*, reflecting relatively lesser effects on RNA structure compared to the former variants as depicted in Figure 6.

### 3.8 Evolutionary Conservation of Candidate gene variations

The conservation analysis **(Figure 8)** revealed that several variants, including TSPEAR L219P, WNT10A A135S, WNT10A V145M, and EDA R156H, are located in highly conserved regions with a conservation score of 9 out of 9, indicating strong evolutionary stability. TSPEAR L219P showed a slightly lower conservation score of 8. Variants such as PAX9 Q145* and TSPEAR I419fs*150 lead to truncations or frameshifts, resulting in the absence of significant portions of the protein. Notably, these disrupted regions consist of amino acids that are highly conserved, suggesting their critical functional importance.

**Figure 8:**
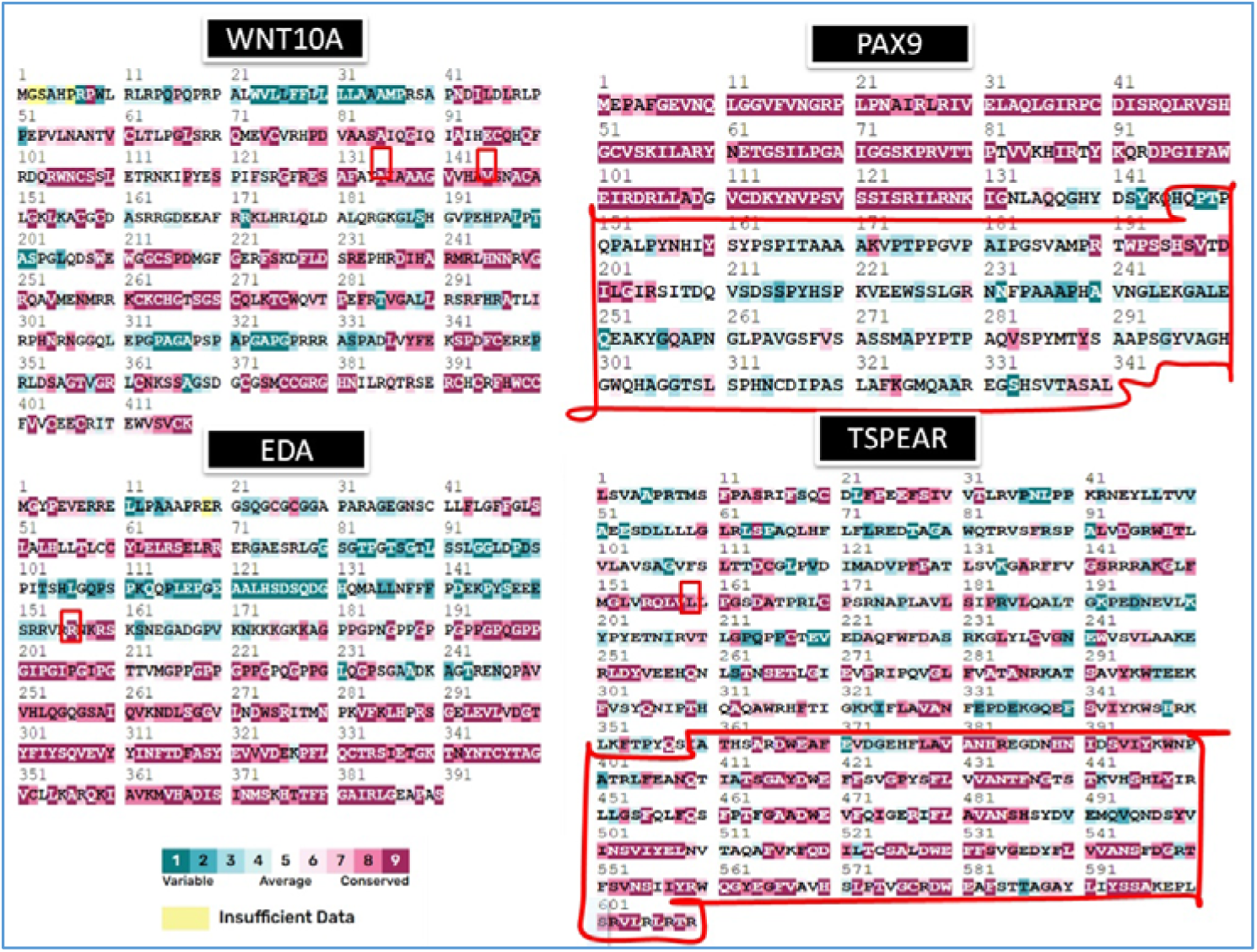
Conservation Analysis of Gene Variants Across WNT10A, PAX9, EDA, and TSPEAR. The figure illustrates the conservation analysis of gene variants across four key genes: WNT10A, PAX9, EDA, and TSPEAR. The sequence alignment highlights regions of variability and conservation using a color-coded scheme. Highly conserved regions are marked in dark pink, indicating sequence stability and potential functional importance. Less conserved or variable regions are shown in shades of blue and teal, with lighter colors reflecting higher variability. Yellow segments indicate areas with insufficient data for reliable analysis. Red boxes outline specific regions containing variants of interest, emphasizing their positions within the protein sequences. This visual representation helps in understanding the evolutionary conservation of these variants, which is crucial for assessing their potential functional impacts and roles in related biological processes.

### 3.9 Protein Structure Stability Analysis

The stability analysis of gene variants (**Table 5)** revealed that TSPEAR L219P, WNT10A A135S, WNT10A V145M, and EDA R156H predominantly show destabilizing effects across multiple prediction tools, indicating a decrease in protein stability. For TSPEAR L219P, all tools consistently predicted a destabilizing impact, with ΔΔG values ranging from -1.284 to -5.93 kcal/mol. WNT10A A135S showed significant destabilization across all models, particularly in DUET (-2.606 kcal/mol). WNT10A V145M had mixed predictions, with most tools indicating destabilization except CUPSAT, which predicted stabilization (0.98 kcal/mol). EDA R156H presented a slight destabilizing trend, with I-Mutant showing the most substantial decrease (-1.20 kcal/mol). Variants TSPEAR I419fs*150 and PAX9 Q145* could not be assessed for stability due to their truncating nature.

**Table 5:** Protein Structure Stability Analysis of Gene Variants.

| Variants | mCSM<br>Predicted<br>Stability<br>Change<br>($\Delta\Delta G$) | SDM<br>Predicted<br>Stability<br>Change<br>($\Delta\Delta G$) | DUET Predicted<br>Stability<br>Change ($\Delta\Delta G$) | CUPSAT | I-Mutant | MUpro |
| --- | --- | --- | --- | --- | --- | --- |
| **TSPEAR:** L219P | -1.284<br>kcal/mol<br>(Destabilizing<br>) | -4.31 kcal/mol<br>(Destabilizing<br>) | -1.959 kcal/mol<br>(Destabilizing) | (Destabilizing<br>) -5.93<br>kcal/mol | -1.38<br>kcal/mol<br>Destabilisin<br>g | -1.8029236<br>(DECREAS<br>E stability) |
| **TSPEAR:**I419fs*150 | NA | NA | NA | NA | NA | NA |
| **PAX9:** Q145* | NA | NA | NA | NA | NA | NA |
| **WNT10A** :A135S | -2.386<br>kcal/mol<br>(Destabilizing<br>) | -2.57 kcal/mol<br>(Destabilizing<br>) | -2.606 kcal/mol<br>(Destabilizing) | Destabilising -<br>2.92 kcal/mol | -0.07<br>kcal/mol<br>Destabilisin<br>g | -0.57849674<br>kcal/mol<br>(DECREAS<br>E stability) |
| **WNT10A:** V145M | -1.387<br>kcal/mol<br>(Destabilizing<br>) | -0.63 kcal/mol<br>(Destabilizing<br>) | -1.454 kcal/mol<br>(Destabilizing) | Stabilising<br>0.98 kcal/mol | -1.31<br>kcal/mol<br>Destabilisin<br>g | -0.69678663<br>kcal/mol<br>(DECREAS<br>E stability) |
| **EDA: R156H** | -0.631<br>kcal/mol<br>(Destabilizing<br>) | 0.17 kcal/mol<br>(Stabilizing) | -0.519 kcal/mol<br>(Destabilizing) | Destabilising<br>-0.83 | -1.20<br>kcal/mol<br>Destabilisin<br>g | -1.5988524<br>(DECREAS<br>E stability) |

**Note:** Negative $\Delta\Delta G$ values indicate destabilizing effects, potentially impacting protein structure. Most tools predict destabilization for variants like TSPEAR: L219P and WNT10A: A135S, with WNT10A: V145M showing mixed results. "NA" denotes unavailable data.

### 3.10 Structural and Functional Analysis of Variants

The tertiary structure analysis showed stability in the case of non-synonymous variants. However, for TSPEAR, the structural comparison of compound heterozygous variants with distance matrix plot, as shown in Figure 9, revealed notable alterations. The secondary structure analysis (Figure 10) revealed structural alterations across the WNT10A, TSPEAR, and EDA genes due to specific mutations. In WNT10A, the novel variant p.A135S and the known variant p.V145M both showed changes in helices and strands compared to the wild-type, suggesting potential impacts on the protein’s stability and function. For TSPEAR, the variant p.L219P and the frameshift mutation p.Ile419Lfs*150 indicated disruptions in transmembrane helices and signal peptides, which may affect protein localization and signaling. In EDA, the p.R156H variant showed alterations in metal-binding and extracellular domains, pointing to possible functional changes. These structural differences highlight how mutations can affect protein conformation and biological activity.

**Figure 9:**
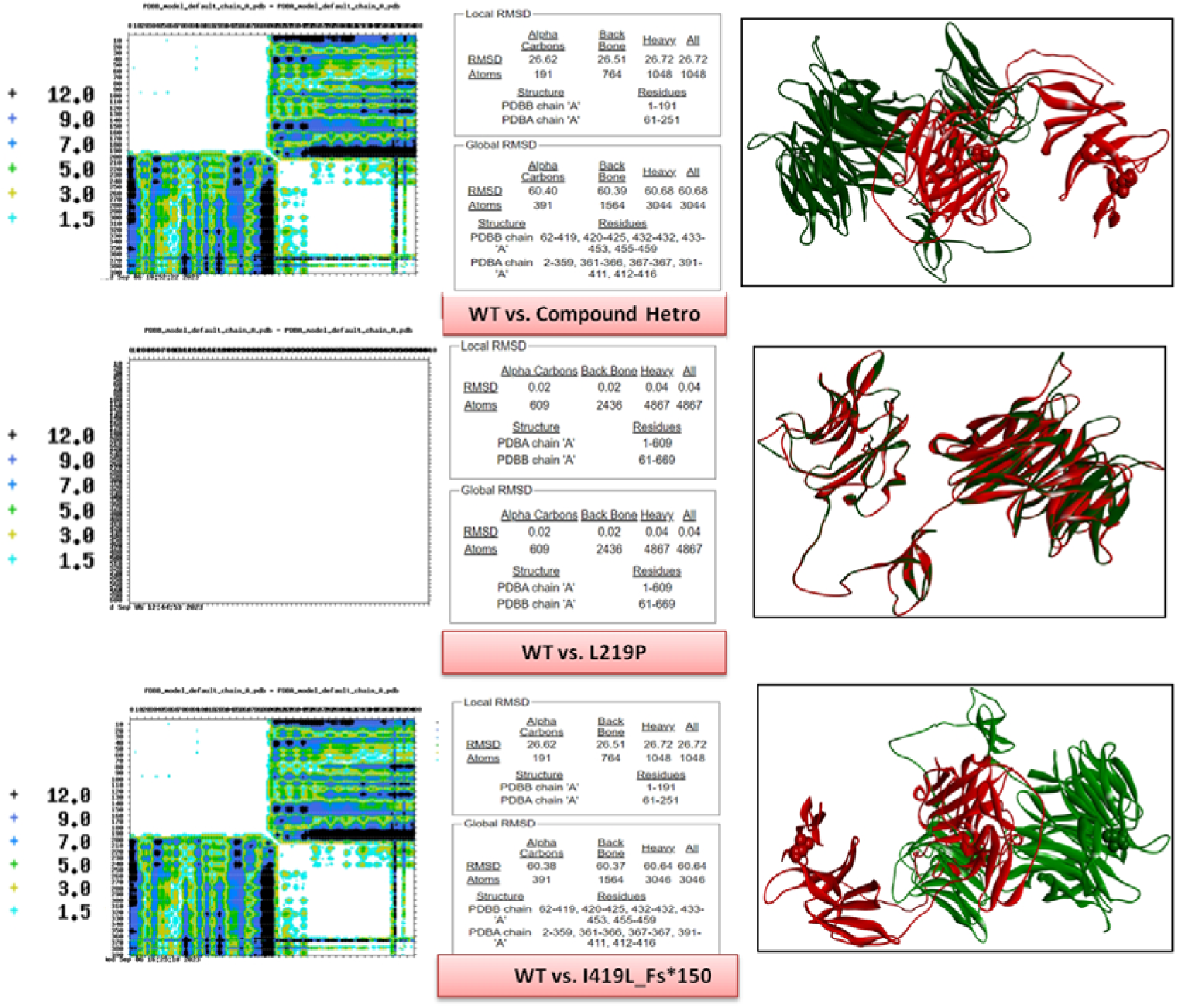
Comparative Analysis of TSPEAR Variants Through Matrix Plot and 3D Structural Evaluation Left Panel: Matrix plot comparing wild-type (WT) and mutant (compound heterozygous and L219P) variants of TSPEAR. Color intensity indicates the extent of structural alterations, with darker shades representing greater structural changes and lighter shades indicating minimal or no changes. Middle Panel: Local and global RMSD (Root Mean Square Deviation) values for the full-length protein, comparing structural deviations across wild-type and mutant variants. Right Panel: 3D structural representations of the TSPEAR variants, with color-coding to denote regions of significant structural differences. Wild-type regions are shown in green, while mutations are highlighted in red. These analyses underscore the impact of mutations on protein stability, flexibility, and interactions, offering insights into the potential functional consequences of these variants.

**Figure 10:**
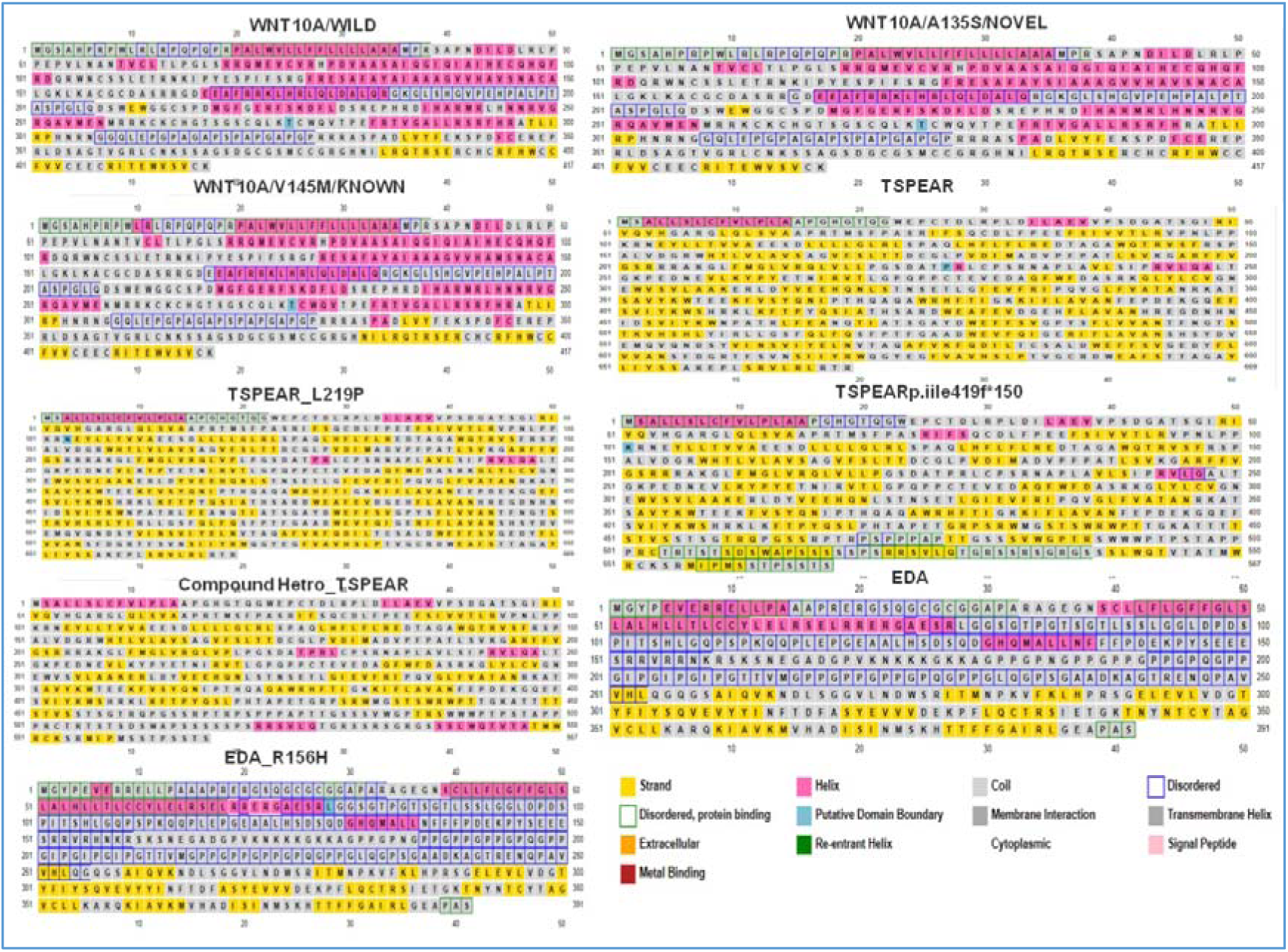
Secondary Structure Prediction of Variants in WNT10A, TSPEAR, and EDA. The figure illustrates the secondary structure prediction of protein variants in the WNT10A, TSPEAR, and EDA genes, highlighting alterations caused by specific mutations. **WNT10A Variants:** Comparisons between wild-type (WILD), known variant (p.V145M), and novel variant (p.A135S). Structural elements like helices, strands, and metal-binding regions are shown, with changes indicating potential structural impact of the variants. **TSPEAR Variants:** Displays predicted structures for p.L219P, compound heterozygous mutations, and a frameshift mutation (p.Ile419Lfs*150). Variants are assessed for alterations in structural domains, including transmembrane helices and signal peptides. **EDA Variants:** Shows secondary structure predictions for the wild-type and p.R156H variant, highlighting potential alterations in metal-binding and extracellular domains. Color coding represents different structural features, such as strands, helices, and metal-binding regions, offering insight into how specific mutations might disrupt or alter protein structure and function.

The protein disorder prediction (Figure 11) analysis showed differences between wild-type and mutant forms across the WNT10A, TSPEAR, and EDA genes. For WNT10A, both the known variant (p.V145M) and the novel variant (p.A135S) exhibited alterations in disordered regions. TSPEAR variants, including p.L219P and the frameshift mutation (p.Ile419Lfs*150), showed shifts in disorder profiles. In EDA, the p.R156H mutant displayed changes in disordered segments.

**Figure 11:**
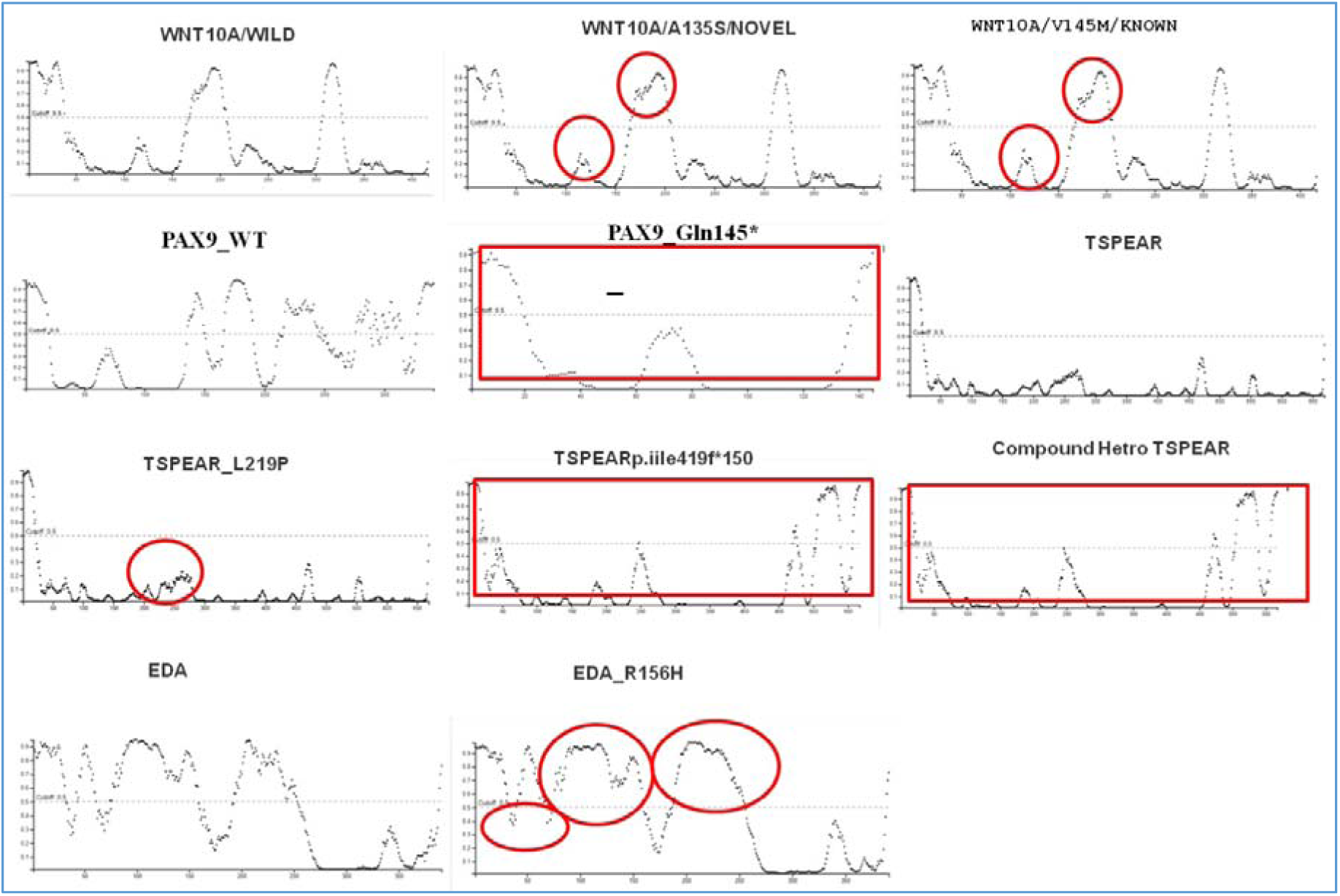
Protein Disorder Prediction of Variants in WNT10A, TSPEAR, and EDA. The figure presents the protein disorder prediction analysis for variants in the WNT10A, TSPEAR, and EDA genes, comparing structural changes between wild-type and mutant forms. **WNT10A Variants:** Disorder profiles for the wild-type, known variant (p.V145M), and novel variant (p.A135S) are depicted. Changes in disorder regions may suggest alterations in protein flexibility or functional sites. **TSPEAR Variants:** Disorder analysis includes p.L219P, compound heterozygous mutations, and frameshift mutation (p.Ile419Lfs*150). Variations in disordered regions highlight the potential impact on protein stability and interaction domains. **EDA Variants:** Displays predicted disorder patterns for wild-type and p.R156H mutant. Differences in disordered segments may affect protein functionality and binding capabilities.

The GO enrichment analysis of the variants (Figure 12-15), compared to the wild type, revealed changes in the functional roles of the affected proteins. Variants showed alterations in biological processes, molecular functions, and cellular components, indicating shifts in protein behavior and potential impacts on associated pathways. PTM analysis (Figure 16) revealed functional alterations in TSPEAR (p.I419F), compound heterozygous TSPEAR, WNT10A (p.A135S), PAX9 (p.Q145*), and EDA (p.R156H) variants. Subcellular localisation analysis (Figure 17) indicated that most variants exhibit similar localization patterns to the wild-type. However, TSPEAR variants displayed distinct shifts, suggesting potential impacts on functionality.

**Figure 12:**
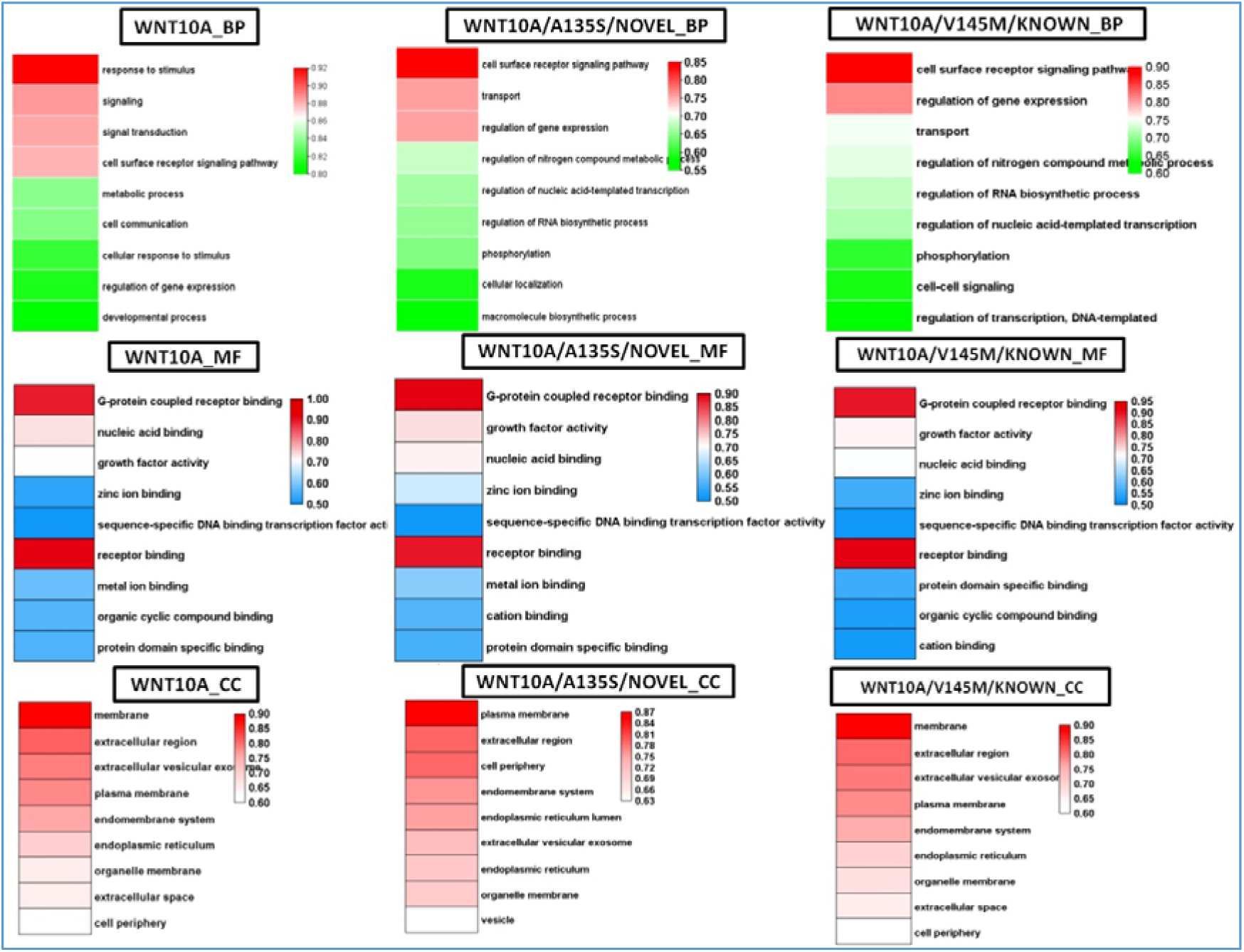
GO enrichment analysis of WNT10A variants. The figure displays the GO enrichment results for three biological processes (BP), molecular functions (MF), and cellular components (CC) associated with WNT10A wild-type (WT), A135S, and V145M variants. The heatmap represents the enrichment score (y-axis) for each GO term (x-axis). The color intensity indicates the significance of the enrichment, with red representing higher significance and blue representing lower significance. The results highlight the functional differences among the WNT10A variants, particularly in terms of their involvement in signaling pathways, cell processes, and molecular functions.

**Figure 13:**
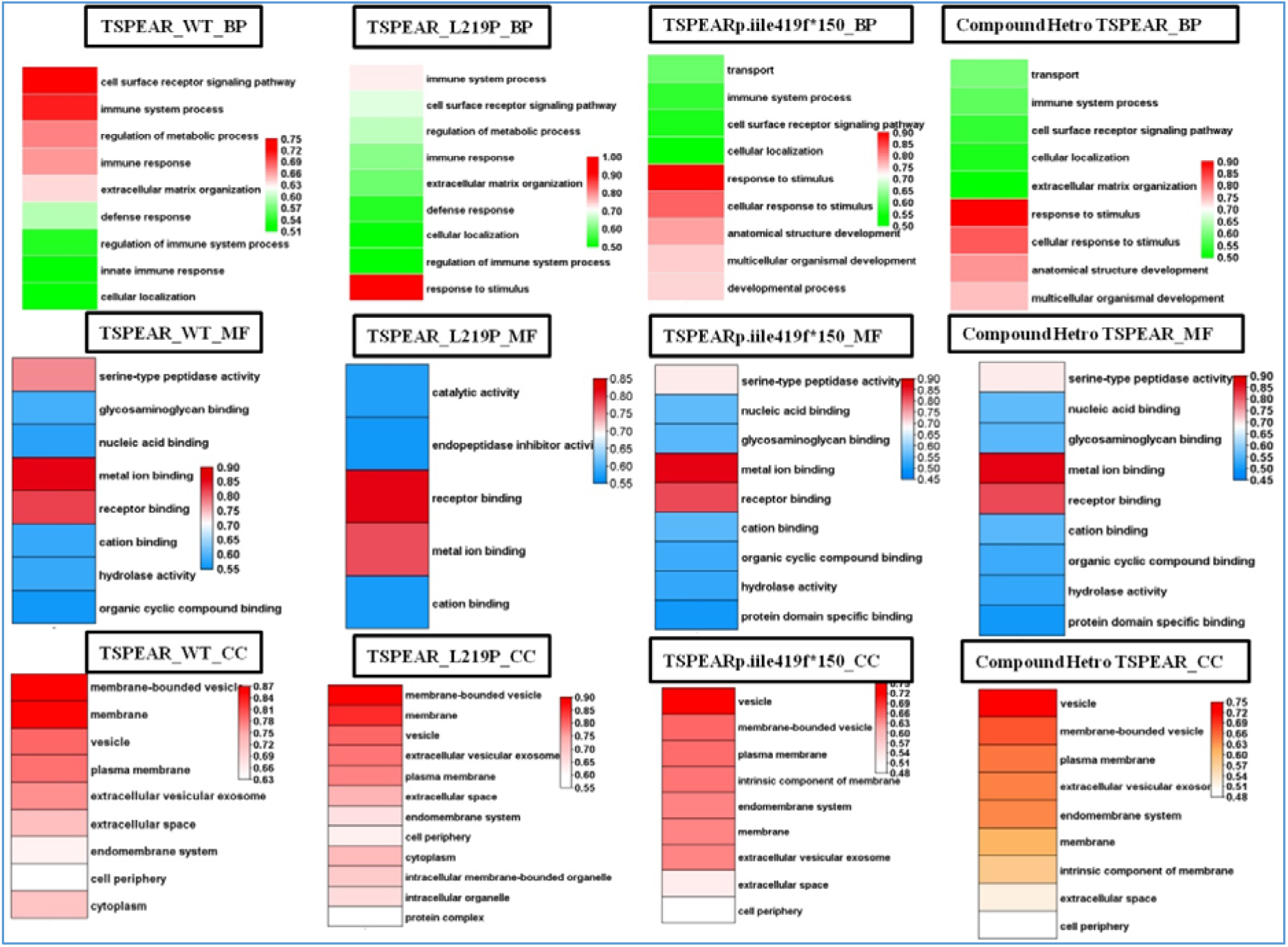
Gene Ontology (GO) enrichment analysis of TSPEAR variants. The figure displays the GO enrichment results for three biological processes (BP), molecular functions (MF), and cellular components (CC) associated with TSPEAR wild-type (WT), L219P, I419f*150 variants, and their compound heterozygous state. The heatmap represents the enrichment score (y-axis) for each GO term (x-axis). The color intensity indicates the significance of the enrichment, with red representing higher significance and blue representing lower significance. The results highlight the functional differences among the TSPEAR variants, particularly in terms of their involvement in immune system processes, cell surface receptor signaling, and extracellular matrix organization.

**Figure 14:**
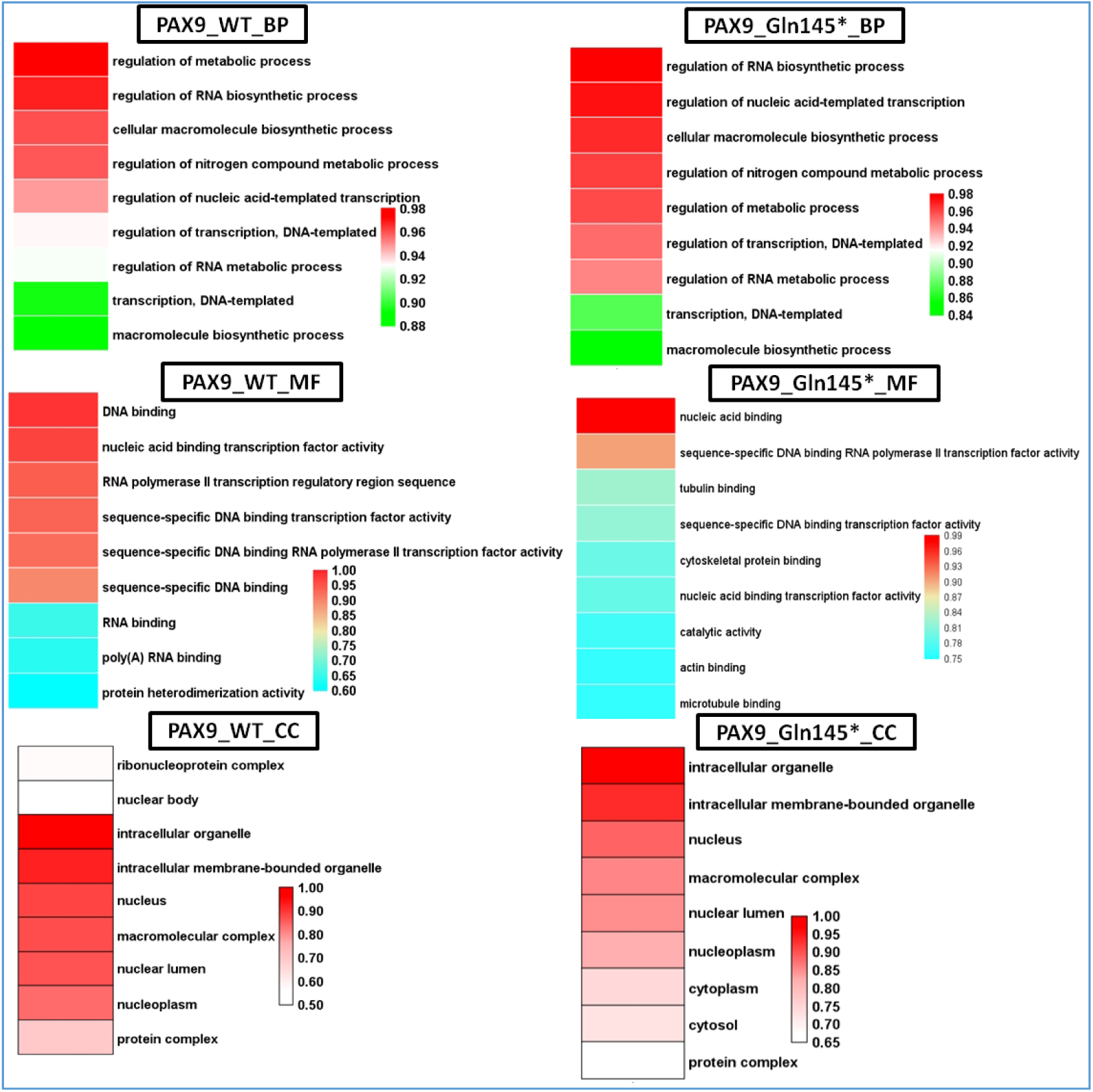
GO enrichment analysis of PAX9 variants. The figure displays the GO enrichment results for three biological processes (BP), molecular functions (MF), and cellular components (CC) associated with PAX9 wild-type (WT) and Gln145* variant. The heatmap represents the enrichment score (y-axis) for each GO term (x-axis). The color intensity indicates the significance of the enrichment, with red representing higher significance and blue representing lower significance. The results highlight the functional differences between the PAX9 variants, particularly in terms of their involvement in gene expression regulation, transcription, and nuclear processes.

**Figure 15:**
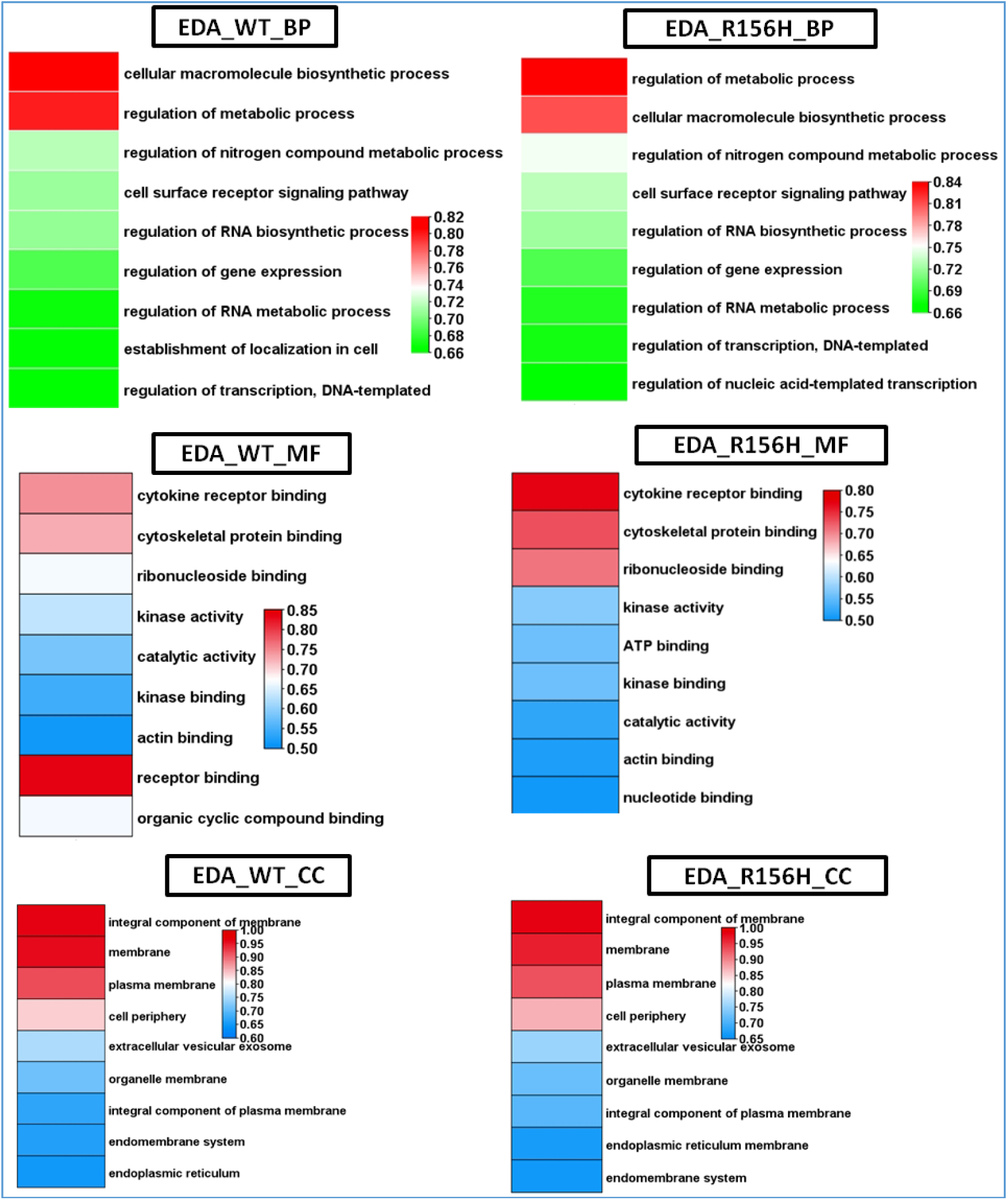
GO enrichment analysis of EDA variants. The figure displays the GO enrichment results for three biological processes (BP), molecular functions (MF), and cellular components (CC) associated with EDA wild-type (WT) and R156H variant. The heatmap represents the enrichment score (y-axis) for each GO term (x-axis). The color intensity indicates the significance of the enrichment, with red representing higher significance and blue representing lower significance. The results highlight the functional differences between the EDA variants, particularly in terms of their involvement in cell signaling, receptor binding, and membrane components.

**Figure 16:**
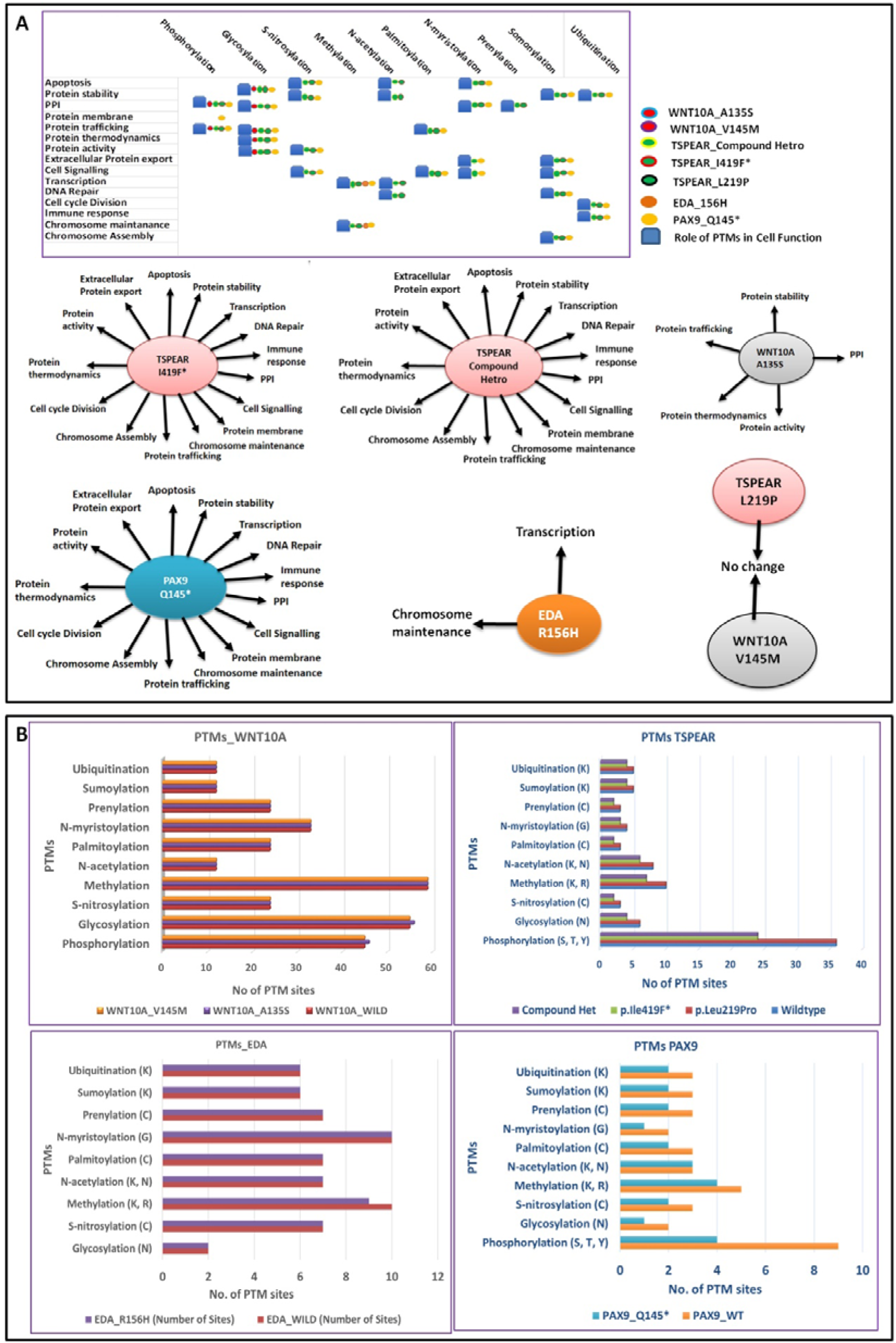
Post-Translational Modifications (PTMs) Enrichment Analysis of WNT10A, EDA, PAX9, and TSPEAR Variants. The figure illustrates the enrichment patterns of different PTMs (such as phosphorylation, ubiquitination, acetylation, etc.) associated with the wild-type (WT) and mutant forms of these genes. **(A)** Highlights the changes in PTMs observed in the variants and their potential functional impacts. **(B)** Shows the fluctuations in PTM levels, comparing the wild-type to the mutant variants, emphasizing variations in PTM profiles.

**Figure 17:**
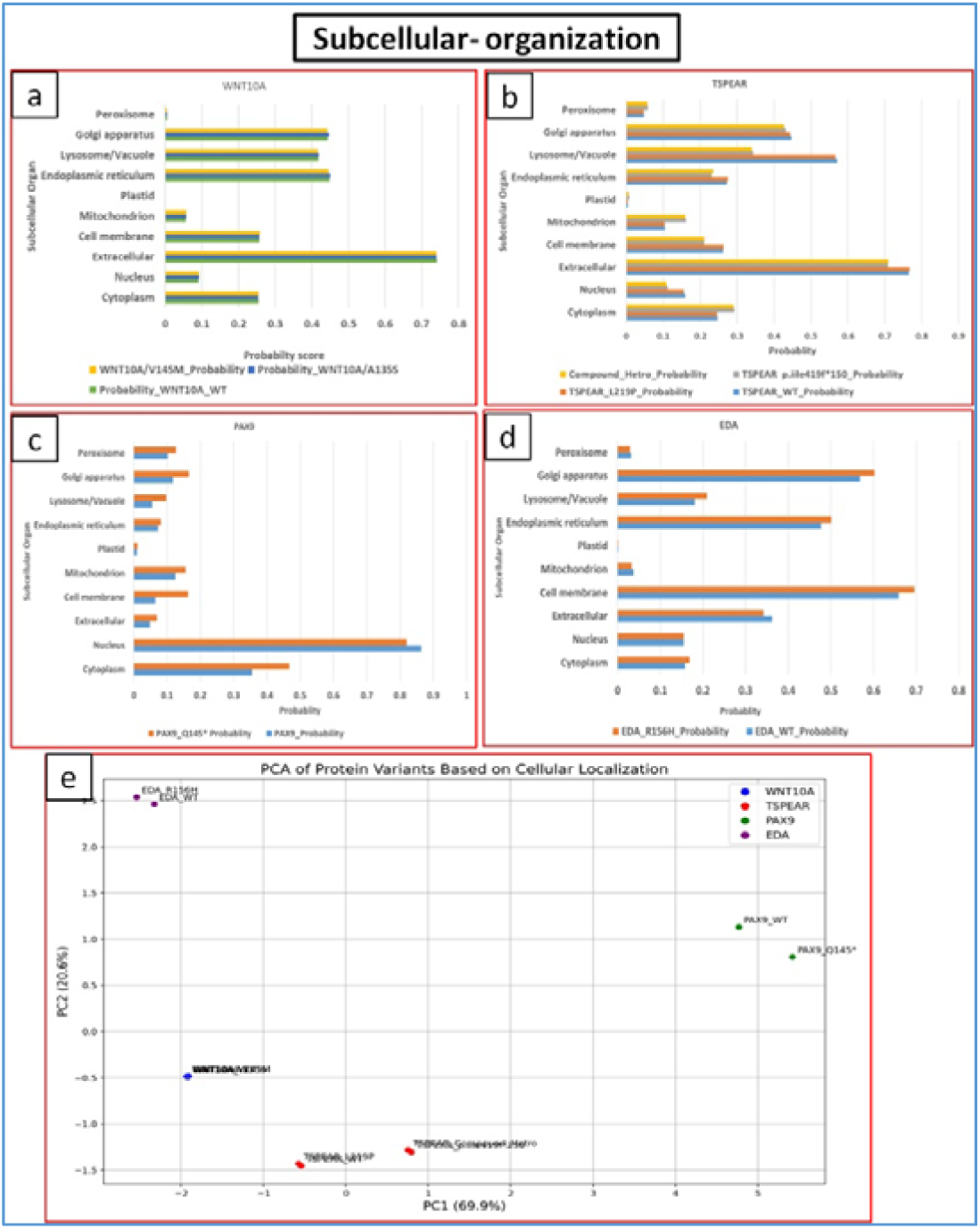
Subcellular localization analysis of WNT10A, EDA, PAX9, and TSPEAR variants. The figure depicts the predicted subcellular localization probabilities for each variant across various organelles (a-d) (peroxisome, Golgi apparatus, lysosome/vacuole, endoplasmic reticulum, plastid, mitochondrion, cell membrane, extracellular space, nucleus, and cytoplasm). (e) The PCA plot illustrates the clustering of protein variants based on cellular localization profiles. EDA wild-type and R156H variant cluster closely, indicating minimal changes in localization. PAX9 wild-type and Q145* variant show slight shifts, suggesting potential alterations. WNT10A wild-type and p.V145M variant cluster together, reflecting little change. TSPEAR variants, including the compound heterozygous and L219P, display distinct positions, indicating significant differences in cellular localization. Overall, most variants show similar localization patterns to wild-type, except for notable shifts in TSPEAR variants, which may affect functionality.

Hydrophobicity changes (Supplementary Information1) among protein variants. WNT10A variants (p.A135S, p.V145M) have slight decreases compared to wild type. PAX9 p.Q145* displays increased hydrophobicity. TSPEAR variants (p.L219P, compound heterozygous) show variable changes. EDA p.R156H has a marked increase, suggesting altered structural properties.

### 3.11 Molecular Simulation Dynamics Analysis

The molecular dynamics simulations **(Figure 18-22)** conducted over a period of 10 ns revealed that the protein variants reached equilibrium, providing valuable insights into their structural stability and flexibility. RMSD showed the highest deviation in EDA_R156H (3.073 Å), while WNT10A_WT had the lowest (0.128 Å), indicating structural stability. RMSF was highest in EDA_R156H (1.106 Å) and TSPEAR_Compound_Hetero (0.675 Å), suggesting increased flexibility. SAS showed a significant increase in EDA_R156H (124.049 Å²), while WNT10A variants remained stable. H-bond counts were lower in EDA and TSPEAR variants, with TSPEAR_Compound_Hetero at 250.722 compared to TSPEAR_WT (421.84). Rg was highest for TSPEAR_Compound_Hetero (2.349 Å), indicating an expanded structure. The comparative analysis of protein dynamics revealed distinct clustering of variants. Variants plotted close to each other on the PCA plot **(Figure 22)** indicated similar biophysical characteristics, while those positioned further apart suggested significant differences due to mutations. Notable insights from the PCA plot included the clustering of WNT10A wild-type and its variants (V145M, A135S), which indicated minimal changes in their biophysical properties. In contrast, TSPEAR variants, including TSPEAR_419fs150 and TSPEAR+ Compound Hetero, were distinctly separated from TSPEAR_WT, suggesting significant alterations in their profiles due to mutations. Additionally, the EDA_R156H and PAX9_WT variants displayed greater dispersion compared to their wild-type counterparts, indicating potential structural or functional changes resulting from the mutations. The overall wider spread of points across the plot highlighted the diversity of the variants, with isolated points, such as EDA_R156H, likely indicating substantial changes in structure or dynamics. Matplotlib, providing a clear representation of the dynamic properties of the simulated variants **(Figure 22)**.

**Figure 18:**
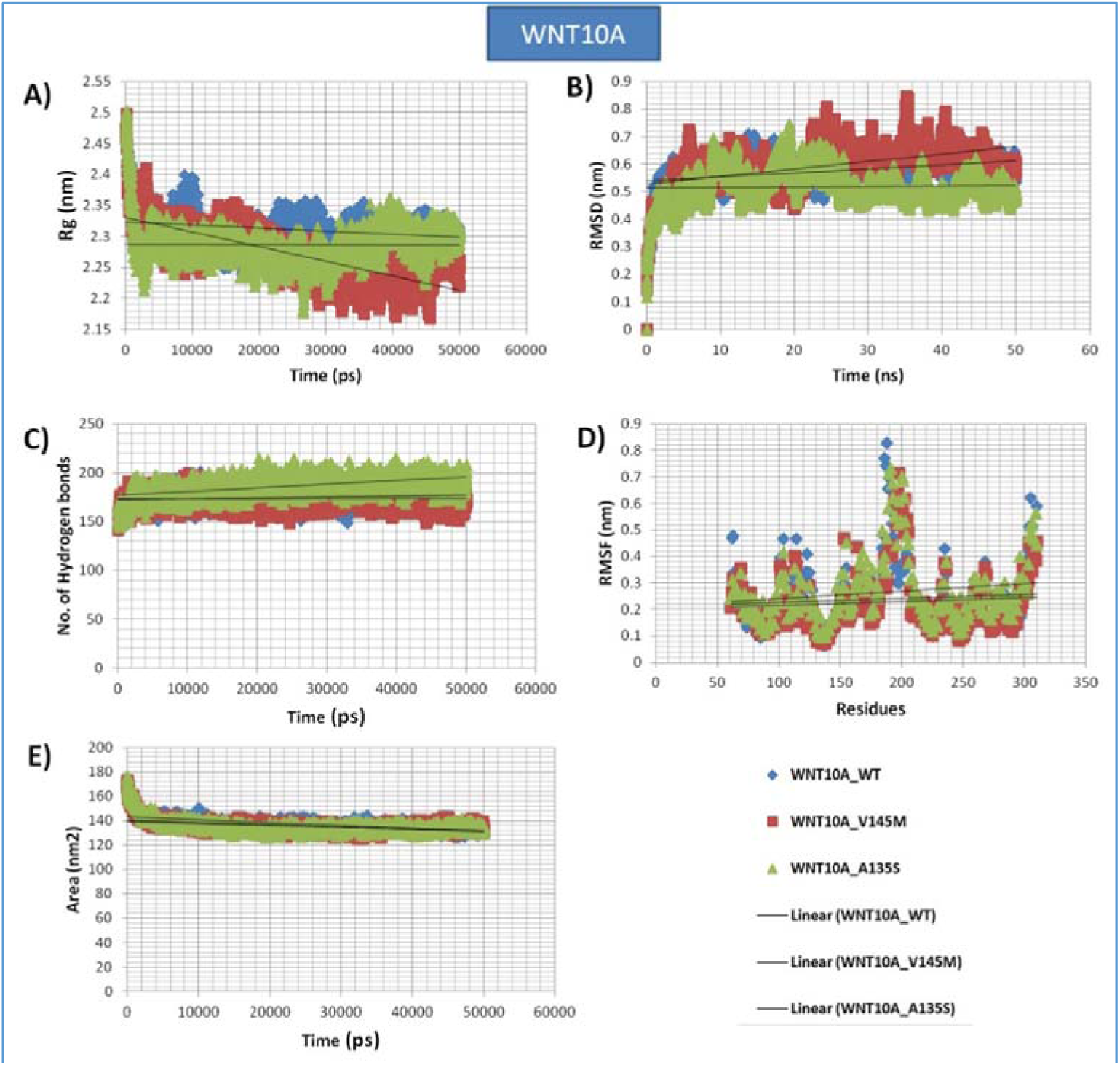
Molecular dynamics simulation analysis of WNT10A variants. The figure depicts the time evolution of various structural and dynamic properties for wild-type (WT) and mutant WNT10A proteins (V145M and A135S). **A:** Radius of gyration (Rg) over time. The graph shows the Rg values over time for WNT10A wild-type (WT), V145M, and A135S variants. The Rg values indicate the compactness of the protein structure, where consistent lower values suggest a more compact structure. WT (blue) shows relatively stable compactness, while variants exhibit fluctuations, with V145M (red) being less stable than A135S (green). **B:** Root-mean-square deviation (RMSD) over time. This panel illustrates the RMSD of the protein backbone over time, reflecting structural stability. WT (blue) maintains lower RMSD, indicating higher stability, whereas V145M (red) shows increased deviation, suggesting structural alterations and reduced stability. A135S (green) remains moderately stable. **C:** Number of hydrogen bonds over time. The number of hydrogen bonds over time is shown for each variant. WT (blue) maintains a higher number of hydrogen bonds, suggesting more stable intramolecular interactions. The variants V145M (red) and A135S (green) show variations, with V145M forming fewer hydrogen bonds, potentially leading to decreased structural stability. **D:** Root-mean-square fluctuation (RMSF) per residue. RMSF values across residues reveal regions of high flexibility and fluctuation. WT (blue) shows lower fluctuations overall, indicating stable regions, while variants, especially V145M (red), show increased fluctuation at certain residues, pointing to local instability. **E:** SAS over time. The SAS values are plotted over time, where lower values indicate reduced exposure to the solvent. WT (blue) shows consistent lower SAS, suggesting a stable and compact structure. V145M (red) has a higher SAS, reflecting more exposure, while A135S (green) shows intermediate values.

**Figure 19:**
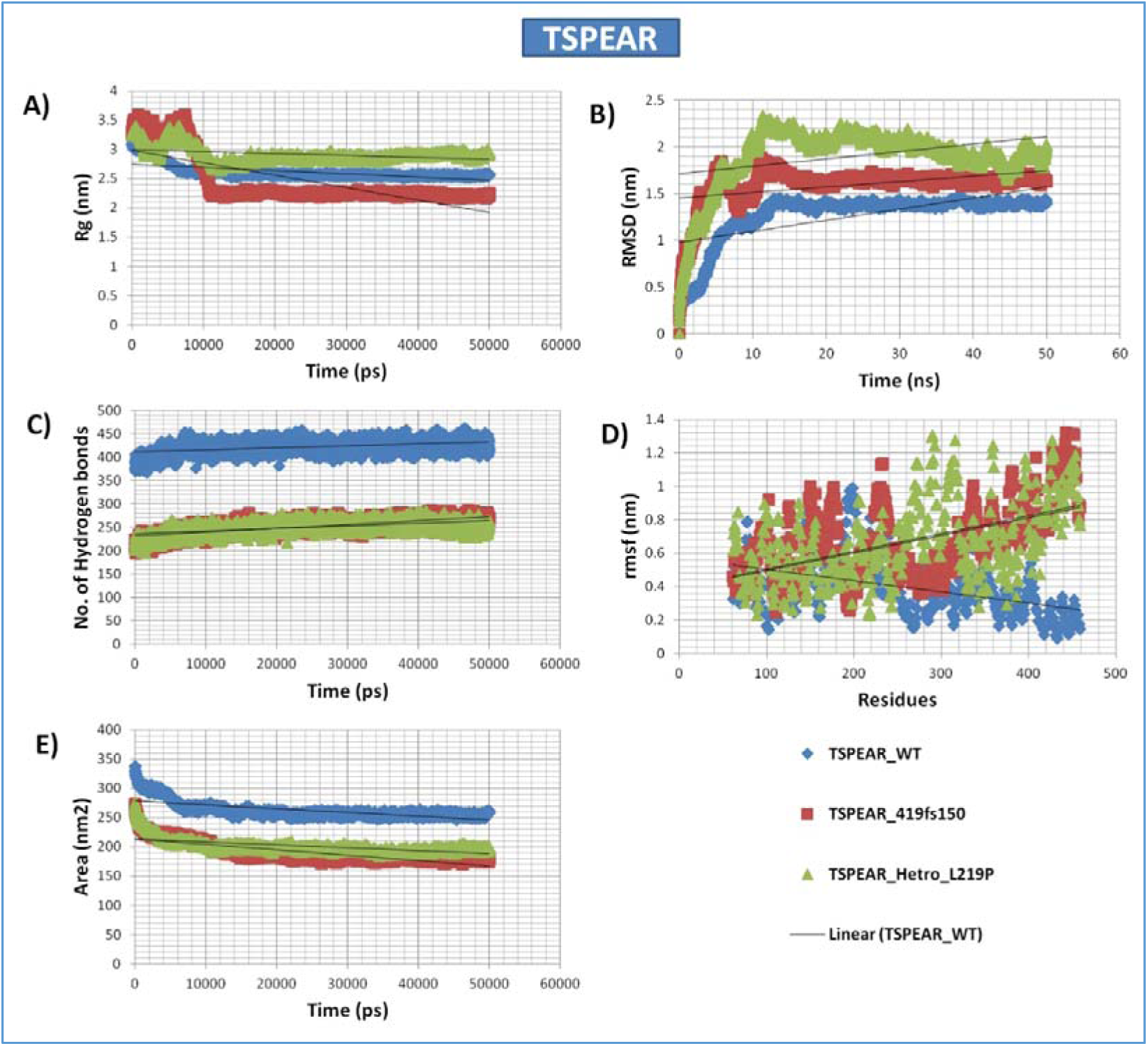
Molecular dynamics simulation analysis of TSPEAR variants. The figure depicts the time evolution of various structural and dynamic properties for wild-type (WT), 419fs150, and L219P variants of TSPEAR. **(A) Radius of Gyration (Rg):** This graph represents the Rg values for TSPEAR wild-type (WT), 419fs150, and Hetro_L219P variants over time, indicating the compactness of each protein structure. The WT (blue) shows a stable Rg, reflecting a consistent compact structure. The 419fs150 variant (red) displays slight fluctuations, while Hetro_L219P (green) shows more variability, suggesting a less compact and potentially more flexible structure. **(B) Root Mean Square Deviation (RMSD):** RMSD plots reveal structural stability across time. The WT (blue) maintains lower RMSD, indicating a stable structure throughout the simulation. The 419fs150 variant (red) shows moderate deviations, while Hetro_L219P (green) has higher RMSD values, suggesting increased structural instability and flexibility. **(C) Number of Hydrogen Bonds:** The panel shows the number of hydrogen bonds formed over time. The WT (blue) maintains a higher and consistent number of hydrogen bonds, suggesting strong intramolecular interactions. The 419fs150 variant (red) shows a similar pattern but with slight reductions, while Hetro_L219P (green) displays fewer hydrogen bonds, indicating weaker structural cohesion. **(D) Root Mean Square Fluctuation (RMSF):** RMSF values are displayed for each residue, revealing regions of high flexibility and fluctuation. WT (blue) exhibits lower overall fluctuations, showing stable regions throughout the protein. The 419fs150 variant (red) shows moderate fluctuations, and Hetro_L219P (green) has the highest fluctuations, indicating regions of instability that may impact function. **(E) SAS:** The SAS plot demonstrates the degree of exposure of the protein to the solvent. WT (blue) maintains a relatively lower SAS, reflecting a compact structure. The 419fs150 variant (red) shows slightly higher SAS, and Hetro_L219P (green) exhibits the highest exposure, suggesting more open and less stable structural conformations.

**Figure 20:**
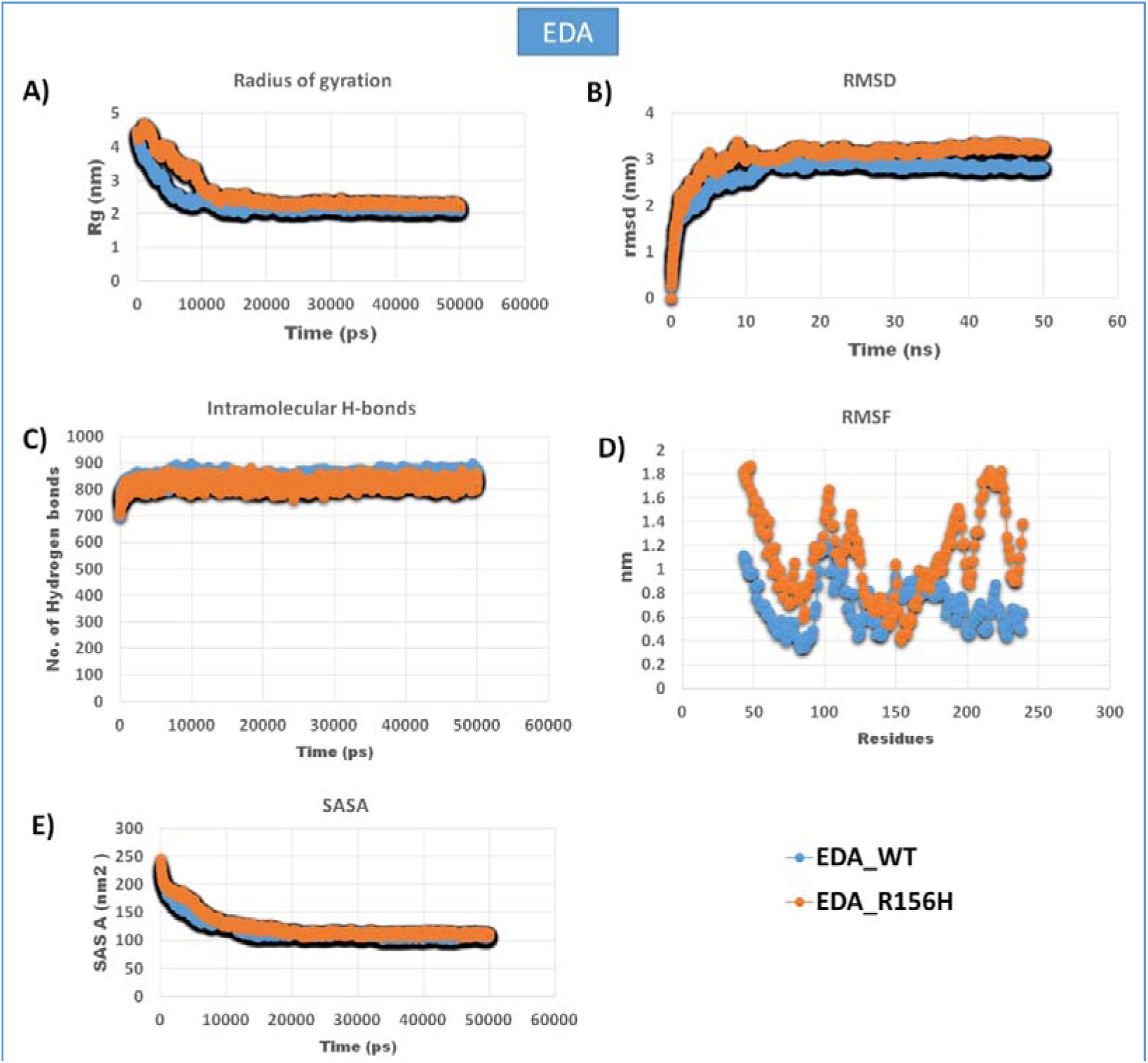
Molecular dynamics simulation analysis of EDA variants. The figure depicts the time evolution of various structural and dynamic properties for wild-type (WT) and R156H variants of EDA. **A:** Radius of gyration (Rg) over time. **B:** Root-mean-square deviation (RMSD) over time. **C:** Number of hydrogen bonds over time. **D:** Root-mean-square fluctuation (RMSF) per residue. **E:** SAS over time. The different line colors represent the different EDA variants, allowing for comparison of their structural and dynamic behavior. The results highlight potential differences in protein stability, flexibility, and interactions due to the R156H mutation.

**Figure 21:**
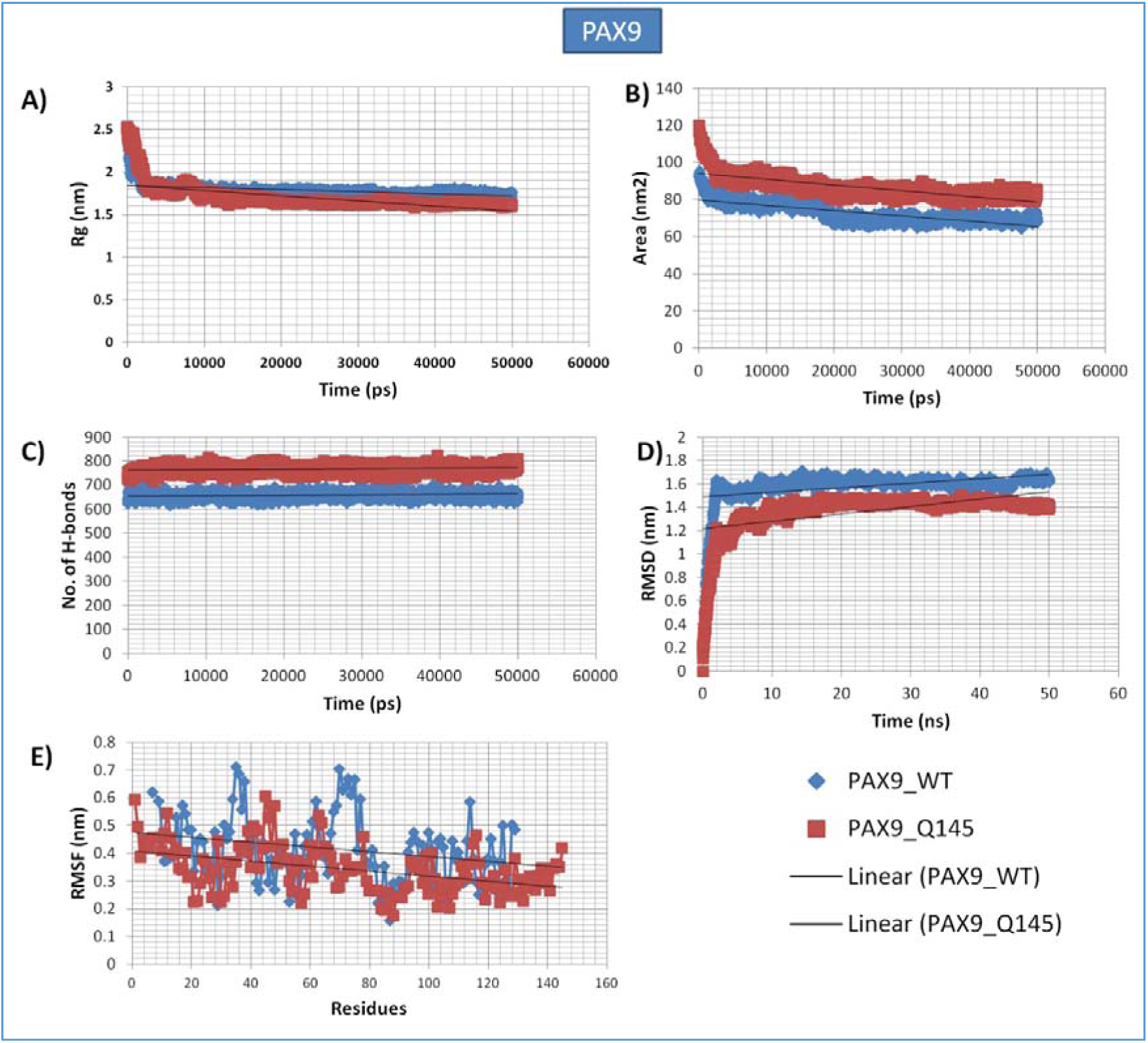
Molecular dynamics simulation analysis of PAX9 variants. The figure depicts the time evolution of various structural and dynamic properties for wild-type (WT) and Q145* variants of PAX9. **A:** Radius of gyration (Rg) over time. **B:** SAS analysis over time. **C:** Number of hydrogen bonds over time. **D:** Root-mean-square deviation (RMSD) over time. **E:** Root-mean-square fluctuation (RMSF) per residue. The different line colors represent the different PAX9 variants, allowing for comparison of their structural and dynamic behavior. The results highlight potential differences in protein stability, flexibility, and interactions due to the Q145* mutation.

**Figure 22:**
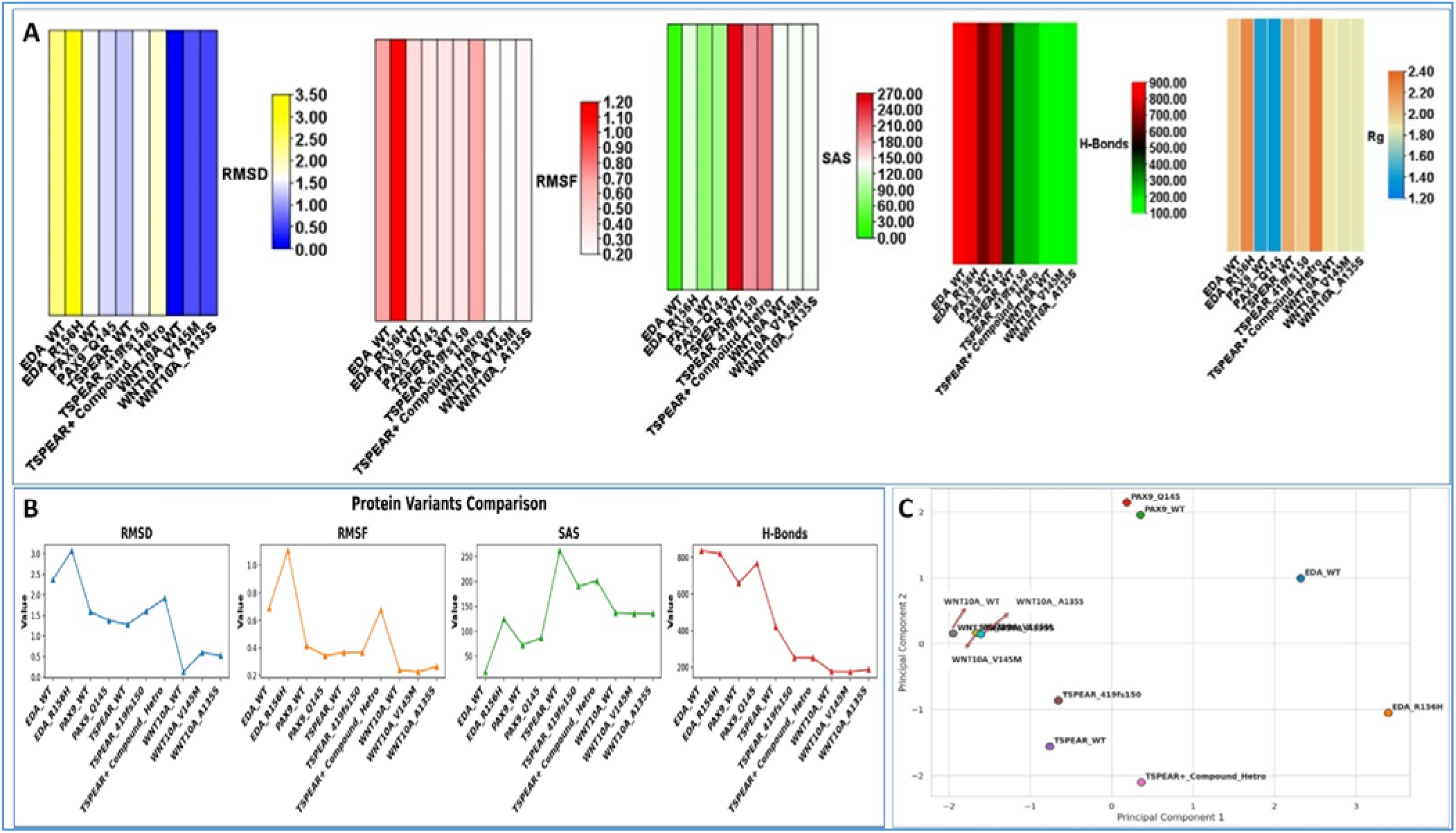
Comparative analysis of protein dynamics and stability for WNT10A, EDA, PAX9, and TSPEAR variants. A. The figure displays the results of molecular dynamics simulations, including: **RMSD:** indicating the overall structural stability of the protein. **RMSF:** revealing the flexibility of different regions within the protein. **SAS**, reflecting the protein’s exposure to solvent. **Rg** measuring the protein’s compactness. **Hydrogen bonds:** The number of hydrogen bonds formed within the protein. The color gradients in each plot represent the different variants, allowing for a visual comparison of their dynamic properties. The results highlight potential differences in protein stability, flexibility, and interactions due to the mutations. B. Matplot for simulated variants. C. Principal Component Analysis (PCA) of Variants and Wild-Type Proteins. The scatter plot shows the PCA results for wild-type (WT) and variant forms of PAX9, WNT10A, TSPEAR, and EDA proteins. Each point represents a different protein or variant, positioned based on their principal component 1 (PC1) and principal component 2 (PC2) scores, indicating the degree of structural variation. Wild-type proteins are labeled with “WT,” while variants are denoted by specific mutations (e.g., PAX9_Q145, EDA_R156H). The plot highlights clustering of WT and mutant proteins, illustrating how structural differences influence their positions in the principal component space, reflecting variations in protein stability, conformation, and functionality. Matplotlib, providing a clear representation of the dynamic properties of the simulated variants.

## 4 Discussion

This study provides an in-depth genetic analysis of congenital tooth agenesis (CTA) by using whole-exome sequencing (WES) and integrating omics data to reveal key genetic variants and their functional impacts. The results identify both known pathogenic variants and novel variants of uncertain significance (VUS), contributing valuable insights into the molecular mechanisms underlying CTA. Significant genetic diversity was observed, with variants identified in key CTA candidate genes, including *WNT10A*, *EDA*, *PAX9*, and *TSPEAR*, among others. This diversity highlights the heterogeneity and complex inheritance patterns characteristic of this condition. Our approach emphasizes the value of WES as a diagnostic and research tool for rare congenital disorders and highlights the functional consequences of specific gene alterations on signaling pathways critical for tooth development.

The WES analysis identified key genetic variants linked to CTA, especially in *WNT10A*, *EDA*, *PAX9*, and *TSPEAR*. This study highlights both known pathogenic and novel variants of uncertain significance (VUS), reflecting the genetic diversity and complex inheritance patterns involved in CTA. Using variant analysis tools and databases like OMIM, EXAC, and gnomAD, the pathogenicity of these variants was consistently supported, providing valuable insights into CTA’s genetic basis.

In *WNT10A*, a homozygous V145M (c.433G>A) variant, classified as likely pathogenic, underscores the critical role of WNT signaling in tooth development. *WNT10A* has previously been associated with several ectodermal dysplasia syndromes that manifest with tooth agenesis, highlighting its influence on ectodermal organogenesis (Bohring et al., 2009). This autosomal recessive (AR) variant aligns with previous studies indicating that *WNT10A* homozygous mutations often lead to more severe phenotypes, such as oligodontia and hypodontia (van den Boogaard et al., 2012). Additionally, the heterozygous novel variant A135S (c.403G>T), classified as a VUS with autosomal dominant (AD) inheritance, introduces a potential new dimension to the inheritance pattern of *WNT10A*-related agenesis. The presence of this variant as a VUS suggests possible gene-environment interactions or oligogenic contributions that may modify its penetrance and expressivity in CTA phenotypes.

The *EDA* gene variant R156H (c.467G>A), found in a homozygous form and linked to X-linked recessive (XLR) inheritance, is classified as likely pathogenic and highlights the role of ectodysplasin A signaling in ectodermal tissue development. Mutations in *EDA* are well-documented in ectodermal dysplasias, often affecting multiple ectodermal structures, including teeth (Cardoso et al., 2021). This variant’s association with tooth agenesis further validates the role of *EDA* in CTA, with consistent damaging predictions across SIFT, Polyphen-2, and MutationTaster2 indicating a probable disruptive effect on EDA signaling. Since *EDA* mutations typically follow XLR inheritance, this variant could contribute to CTA, particularly in male patients who are hemizygous for the mutation.

The *PAX9* gene, which encodes a transcription factor crucial for tooth development, also exhibited two variants. A stop-gain mutation, Q145* (c.433C>T), classified as pathogenic with AR inheritance, is consistent with prior findings that pathogenic *PAX9* mutations contribute to non-syndromic oligodontia (Bonczek et al., 2017). Additionally, a novel VUS (c.-31C>A), associated with AD inheritance, further complicates the genetic picture in CTA. The AD VUS may suggest a less severe or subclinical phenotype, indicating that not all *PAX9* mutations manifest with complete penetrance in CTA. Such variants emphasize the importance of considering diverse inheritance models when evaluating CTA’s genetic etiology, as heterozygous variants in *PAX9* may also impact dental development in a dose-sensitive manner.

The *TSPEAR* gene cause both Syndromic and non-Syndromic CTA(Du et al., 2018)(Peled et al., 2016). The identification of heterozygous variants in the TSPEAR gene in both affected siblings, with one variant inherited from an unaffected mother, raises critical questions about the gene’s role in disease pathology. TSPEAR is associated with developmental processes, and haploinsufficiency has been linked to phenotypic manifestations in affected individuals. The presence of two variants—L219P (c.656T>C) and a frameshift mutation I419Lfs150 (c.1255delA)—suggests that compound heterozygosity may contribute to the clinical phenotype.

The second variant may be a de novo mutation or be inherited from the father, whereas the first variant is transmitted from the mother. Because the I419Lfs150 frameshift mutation results in a shortened protein, it may alter its function and impact ectodermal structures, which is important for diseases like tooth agenesis. The relevance of assessing both hereditary and de novo mutations in genetic evaluations is highlighted by the possibility that both variations, although being categorized as variants of unknown significance (VUS), may jointly contribute to the phenotype.

In order to clarify these variations’ contributions to CTA and improve our knowledge of the genetic foundation of this disorder, more functional research is required to determine how these variants affect TSPEAR activity and their function in dental epithelial signaling. Further functional validation is necessary to ascertain if the defective TSPEAR protein affects tooth epithelial signaling, since the compound heterozygous presentation implies that both alleles contribute to the phenotype.

All identified variants were assessed across predictive tools, including SIFT, Polyphen-2, MutationTaster2, and CADD3 Phred scores, all of which indicated damaging effects for pathogenic variants in *WNT10A*, *EDA*, *PAX9*, and *TSPEAR*. This consistent pathogenicity prediction across tools and databases strengthens the evidence that these variants likely contribute to CTA. The use of multiple databases, such as OMIM, EXAC, and gnomAD, allowed us to cross-reference allele frequencies and variant classifications, enhancing the robustness of our analysis. Specifically, the presence of these variants in disease-associated databases and their classification as likely damaging through computational models underscores the relevance of these candidate genes in CTA pathogenesis.

The RT-PCR analysis reveals that variants in *EDA* and *WNT10A* significantly reduce the expression levels of these genes in CTA samples. Specifically, *EDA* expression showed marked downregulation, with ΔCT values reflecting substantial decreases relative to the internal control (*p* < 0.0001). Similarly, *WNT10A* expression was also notably diminished in samples carrying the variants (*p* < 0.001), suggesting a direct impact of these mutations on WNT and EDA signaling pathways. This downregulation supports the hypothesis that impaired WNT and EDA signaling contributes to tooth agenesis by disrupting essential pathways in ectodermal tissue development. Prior studies emphasize the roles of *EDA* and *WNT10A* in regulating ectodermal differentiation, further validating their association with phenotypes seen in CTA (Bohring et al., 2009)(Cardoso et al., 2021).

This WES gene panel analysis has identified a curated set of 709 genes with potential relevance to congenital CTA, providing critical insights into the genetic underpinnings of this condition. By filtering variants with a minor allele frequency (MAF) threshold of ≤20% and comparing patient-specific variants against controls, the study narrowed down candidate genes that may influence CTA. The final gene panel includes both rare variants of uncertain significance (VUS) and benign variants, offering a comprehensive view of both potential pathogenic contributors and common polymorphisms in CTA.

Among the 21 genes commonly present in all patient samples, variants in *TM4SF20*, *LCORL, OR4F21*, *TNS2*, *VWA8*, and *MYH6* were identified. These genes contain both synonymous and non-synonymous variants that span benign and VUS classifications. Notably, *OR4F21* exhibited two novel VUS (K310R, F44L), indicating possible regulatory functions yet to be fully understood. The observed variation in *LCORL* (L1734P, P1709R), a gene previously associated with growth disorders and Alzheimer’s, suggests it may play a broader role in developmental pathways relevant to CTA (Scelsi et al., 2018)(Lin et al., 2017). Similarly, TNS2 is involved in kidney diseases and cancer (Ashraf et al., 2018)(Cheng et al., 2018), while VWA8 variants are implicated in kidney diseases, cleft lip and palate, cancer, and autism, suggesting cross-system genetic influences that could indirectly impact tooth development. (Aylward et al., 2016)(Anney et al., 2010)(Guo et al., 2023).

The identified variants across genes like *TM4SF20* (autism and Alzheimer’s) (Krgovic et al., 2022)(Floudas et al., 2014)(Laskowski et al., 2018), *MYH6* (CHD) (Granados-Riveron et al., 2010), and *KNL1* (microcephaly, Cancer) (Genin et al., 2012)(He et al., 2023) highlight potential genetic contributors to systemic or developmental abnormalities, emphasizing the pleiotropic nature of CTA. The presence of two consistently observed variants, *OR4F21* (K310R) and *MRTFB* (A135), highlights genetic diversity across the six CTA cohort. Notably, the *MRTFB* variant has been associated with autism and CHD (Holt et al., 2010), supporting its relevance in neurodevelopmental and cardiovascular disorders. In contrast, *OR4F21* (K310R) has not yet been linked to any specific disease, suggesting it may represent a unique, potentially VUS variant within this cohort. Further investigation could clarify *OR4F21’s* role, if any, in developmental anomalies or other conditions. These findings highlight the importance of exploring shared genetic factors across these conditions, which may contribute to the comorbidity and varied clinical presentations observed in patients. Pathway enrichment analysis of the WES panel demonstrated significant roles in DNA damage response, cell cycle regulation, and apoptotic signaling pathways, all critical to cellular stability and development. The involvement of biological processes like DNA replication and repair aligns with the rapid cell division and differentiation required during early tooth bud formation. Additionally, enriched cellular components, including mitochondrial complexes and cell junctions, imply a substantial role for these genes in energy metabolism, cell communication, and genomic stability—all essential for normal tooth development (Sui et al., 2023)(Nijakowski et al., 2023). The R158Q variant in the *LOX* is classified as benign, with a frequency of 17.02%, suggesting evolutionary neutrality. This c.473G>A nucleotide change leads to a non-synonymous coding mutation that does not impair LOX function. As a top 10 hub gene, LOX plays a critical role in collagen and elastin cross-linking, maintaining extracellular matrix stability. Although benign, the variant’s presence in three patient samples, highlights its relevance and possible involvement in pathways associated with cancer, diabetes, and heart conditions. Further studies could explore any subtle phenotypic effects associated with its prevalence.

PPI network analysis revealed a well-connected structure with 470 nodes and 1211 interactions, including 11 network modules and key hub genes such as *TNF, TP53*, *STAT3*, and *CXCL12*. The clustering of developmental genes within the network suggests their relevance in tooth morphogenesis, with central genes like *TP53* and *STAT3* indicating their involvement in cellular proliferation and apoptosis, both critical to the formation and patterning of teeth (Romano et al., 2012)(Yamashiro et al., 2022). The network’s connectivity metrics, with an average of 5.153 neighbors per node and a clustering coefficient of 0.126, underscore the strong interactions between these candidate genes, emphasizing their potential coordinated role in CTA pathogenesis.

The multi-omics integration analysis, combining data from the WES panel and GSE56486 datasets, highlights several shared genes and pathways potentially involved in CTA. By identifying gene overlaps across WES Panel, GSE56486_UP, and GSE56486_Down datasets, this approach underscores both unique and shared molecular signatures in CTA. The integration revealed 18 common genes between WES and upregulated GSE56486 genes, and 15 between WES and downregulated GSE56486 genes, suggesting functional or regulatory relationships that may influence dental development.

Pathway enrichment of upregulated genes identified associations with NOD-like receptor signaling, steroid biosynthesis, and cytoskeletal metabolism. These pathways have well-established roles in immune response regulation, cellular signaling, and structural stability, suggesting that dysregulation could impact the ectodermal tissues involved in tooth formation (Lopez-Pajares et al., 2013). Conversely, downregulated genes were linked to pathways like endocytosis, bacterial invasion of epithelial cells, and fatty acid degradation, hinting at compromised metabolic activities that could indirectly influence cellular processes essential for tooth morphogenesis.

The functional analysis further emphasized that upregulated genes are primarily associated with binding activities, enzymatic functions, and extracellular matrix formation, which are essential for tissue adhesion and structural integrity during tooth development (Nowwarote et al., 2022). Downregulated genes, meanwhile, are involved in oxidoreductase activities, cytoskeletal binding, and ion transport—processes critical for cellular organization and signaling. Notably, downregulated genes’ involvement in cell junctions and cytoskeleton components may indicate disrupted cellular cohesion or communication in CTA, potentially affecting morphogenesis and cellular architecture.

Among the 33 differentially expressed genes (DEGs), notable variants were identified. *ENAH*, which displayed a benign deletion variant (ERQERLD217D) in multiple samples, is known for its role in actin dynamics and cell motility, which are crucial during morphogenesis (Hwang et al., 2022). Additionally, *LOX* (R158Q), with a benign non-synonymous variant, is involved in extracellular matrix stabilization, further supporting the role of structural integrity in CTA. Several genes, including *IRF5*, *CEL*, and *ABTB2*, presented VUS, adding potential regulatory or structural effects that could contribute to variability in phenotypes. Moreover, benign and VUS variants in genes like *RNF175*, *TGFB1*, and *PRKCB*—which play roles in immune signaling and tissue remodeling—suggest genetic factors that may influence disease progression and severity.

The mutaRNA analysis reveals significant variations in the structural impacts of single nucleotide changes in RNA snippets of the examined genes. Utilizing remuRNA’s computed relative entropy scores (H(wt:mu)), we assess how specific SNPs perturb local RNA structures, with higher scores indicating greater alterations. The *WNT10A* c.403G>T variant stands out with a relative entropy score of 3.409, suggesting substantial structural changes, while *EDA* c.467G>A (2.248) and *TSPEAR* c.656T>C (1.392) also indicate notable impacts. In contrast, *WNT10A* c.433G>A shows a lower score of 0.355, indicating a lesser effect on RNA structure. Base pairing probabilities and accessibility profiles further elucidate these structural consequences (Figures 1, 2, 3). The pronounced alterations in base pairing, particularly for *WNT10A* c.403G>T and *TSPEAR* c.656T>C, suggest disruptions in RNA folding that may affect function and stability. Although *WNT10A* c.433G>A and *EDA* c.467G>A exhibit only minor structural changes, even slight variations can influence RNA-protein interactions, which are crucial for genes like *WNT10A*, involved in development and tissue homeostasis.

Overall, these findings highlight the necessity of considering RNA accessibility alongside structural features to fully understand the functional consequences of genetic variants. This research underscores the importance of further experimental validation and functional assays to clarify the precise effects of these variants on RNA structure and function, providing insights into the molecular mechanisms related to disease pathogenesis or specific cellular phenotypes (Salari et al., 2013).

The evolutionary conservation and protein structure analyses reveal critical insights into how specific gene variants impact protein stability and function, underscoring their potential roles in CTA. The conservation analysis highlighted highly conserved variants, such as *TSPEAR L219P*, *WNT10A A135S*, *WNT10A V145M*, and *EDA R156H*, with conservation scores of 9 out of 9. This strong conservation suggests that these amino acids play essential roles in protein function, where mutations are likely to have significant biological consequences. The slight decrease in conservation for *TSPEAR L219P* (score of 8) still indicates considerable functional importance, while the truncating mutations in *PAX9 Q145* and *TSPEAR I419fs*150 remove highly conserved regions, further supporting their likely disruptive impact.

The structural stability analysis across multiple predictive tools identified *TSPEAR L219P*, *WNT10A A135S*, *WNT10A V145M*, and *EDA R156H* as predominantly destabilizing, suggesting a decrease in protein stability. These destabilizing effects, indicated by negative ΔΔG values, imply structural changes that could impair protein function, especially in *TSPEAR L219P* with ΔΔG values as low as -5.93 kcal/mol. For *WNT10A A135S*, a consistent destabilization was observed across models, particularly in DUET (-2.606 kcal/mol). Mixed predictions for *WNT10A V145M* suggest that some aspects of the protein structure may tolerate the mutation, although overall stability is likely reduced. In contrast, *EDA R156H* shows a modest destabilizing trend with ΔΔG of -1.20 kcal/mol, hinting at a minor impact on stability that may still influence biological function. Truncating variants like *PAX9 Q145* and *TSPEAR I419fs*150 were not evaluated for stability due to their nature but are presumed to have severe functional impacts due to significant protein loss.

The tertiary structure analysis of these gene variants revealed marked structural changes in proteins with non-synonymous variants. For instance, *WNT10A* variants (A135S and V145M) demonstrated alterations in helices and strands, potentially affecting protein folding and function. The structural analysis of *TSPEAR* variants (L219P and I419fs*150) identified disruptions in transmembrane helices and signal peptides, likely influencing protein localization and cell signaling capabilities. These structural disturbances in *TSPEAR* could impair its function in ectodermal tissue development, relevant to CTA pathogenesis. Additionally, *EDA*’s R156H variant was associated with altered metal-binding and extracellular domains, suggesting potential changes in the protein’s extracellular signaling function, critical for ectodermal organ development(Bohring et al., 2009).

Protein disorder predictions further support the destabilizing nature of these variants, with disorder profile changes in the *WNT10A*, *TSPEAR*, and *EDA* genes indicating altered flexibility in mutant forms. Both *WNT10A* variants (A135S, V145M) showed shifts in disordered regions, which could impact protein stability and interactions. The frameshift in *TSPEAR* (I419fs*150) notably shifts disorder profiles, while the *EDA* R156H variant exhibits altered disorder segments, potentially modifying extracellular interactions in pathways crucial for tooth development. The post-translational modifications (PTMs) observed in proteins linked to tooth development, including WNT10A, TSPEAR, EDA, and PAX9, reveal diverse impacts on cellular functions such as protein stability, signaling, and chromosome maintenance (Audagnotto & Dal Peraro, 2017). These mutations affect critical pathways involved in cellular homeostasis and developmental processes essential for tooth formation. The distinct roles of each variant underscore how specific PTM changes can contribute to dental abnormalities, highlighting potential targets for therapeutic intervention in congenital tooth defects. This analysis enhances our understanding of PTM-driven mechanisms in tooth morphogenesis and could inform future strategies in dental tissue engineering.

GO enrichment analysis of these variants relative to wild-type forms showed alterations in biological processes, molecular functions, and cellular components, suggesting that these variants influence protein roles in critical pathways. *WNT10A* variants displayed reduced hydrophobicity, while *PAX9 Q145* exhibited increased hydrophobicity, indicating altered structural properties that may affect protein folding and stability. *TSPEAR* compound heterozygous variants showed variable changes in hydrophobicity, while *EDA R156H* demonstrated a notable increase, which could alter protein conformation and interactions within the cellular environment.

The molecular dynamics (MD) simulations provide a detailed assessment of the structural stability, flexibility, and biophysical properties of protein variants linked to CTA. Over a 10 ns simulation period, proteins reached equilibrium, enabling an analysis of their structural deviations and interactions. Root Mean Square Deviation (RMSD) values indicate that the *EDA*_*R156H* variant experiences the highest deviation (3.073 Å), suggesting considerable structural instability. In contrast, *WNT10A*_*WT* remains highly stable, with an RMSD of just 0.128 Å, reflecting the stability of this protein’s wild-type conformation. RMSF values reveal that *EDA_R156H* and *TSPEAR*_*Compound_Hetero* display the highest flexibility, at 1.106 Å and 0.675 Å, respectively, potentially disrupting specific interactions or affecting protein function critical to signaling pathways in tooth formation. The SAS values show that *EDA_R156H* has a markedly increased surface area (124.049 Å²), which could indicate partial unfolding, thereby altering interactions with other proteins or ligands. Hydrogen bond counts are notably lower in *EDA* and *TSPEAR* variants; *TSPEAR_Compound_Hetero*, for example, has only 250.722 H-bonds compared to *TSPEAR_WT* with 421.84, suggesting reduced intra-molecular stability due to mutations. The Radius of Gyration (Rg) data reveals that *TSPEAR_Compound_Hetero* has the highest Rg value (2.349 Å), indicating an expanded structure that may impact the protein’s stability and localization. Structural expansion, especially in proteins with signaling functions, could affect their proper localization and interactions within ectodermal tissues relevant to dental development. PCA results offer a comparative view of the biophysical characteristics of these variants. *WNT10A* wild-type and its variants (V145M, A135S) cluster closely, suggesting minimal structural alteration. Conversely, *TSPEAR* variants, including *TSPEAR_419fs150* and *TSPEAR+Compound Hetero*, are distinctly separated from *TSPEAR_WT*, highlighting significant structural changes. Notably, the *EDA_R156H* variant showed substantial dispersion from its wild type, reinforcing its likely structural or functional impact in CTA.

## 5. Conclusion

This study reveals key genetic variants associated with CTA through WES and multi-omics integration. It significantly enhances our understanding of the genetic underpinnings of CTA by uncovering both novel and established variants in crucial genes involved in tooth development. The integrative multi-omics analysis reveals the intricate nature of CTA and underscores the power of these approaches in dissecting its complex genetic landscape. Key genes such as *WNT10A*, *EDA*, *PAX9*, and *TSPEAR* have emerged as central contributors to the pathogenesis of tooth agenesis, offering valuable insights for future functional investigations. Disruption in critical signaling pathways, including WNT and EDA, plays a significant role in tooth development, as revealed by functional analyses. Additionally, in silico functional and structural predictions provide further support for their involvement in the underlying mechanisms of tooth agenesis, paving the way for deeper exploration in future research. The identified genetic variants also show associations with a range of systemic conditions, including AZ, ALS, autism, CHD, nephropathy, cleft lip palate, COVID-19, and cancer. For instance, *TM4SF20* is linked to AZ and autism, while *LCORL* is implicated in growth disorders and AZ. *TNS2* is associated with kidney diseases, and *MYH6* with CHD. Additionally, *VWA8* is connected to both COVID-19 and autism, and *MRTFB* is involved in both autism and CHD. These findings suggest potential shared genetic pathways between tooth agenesis and various systemic conditions. Moving forward, further research is essential to validate the identified VUS and explore their functional roles in developmental signaling pathways, which could ultimately inform therapeutic strategies for CTA and related disorders.

## Abbreviations

CTA: Congenital Tooth Agenesis
WES: Whole Exome Sequencing
GWAS: Genome-Wide Association Studies
OMIM: Online Mendelian Inheritance in Man
MAF: Minor Allele Frequency
qRT-PCR: Quantitative Reverse Transcription Polymerase Chain Reaction
CCDS: Consensus Coding Sequence
VUS: Variants of Uncertain Significance
AR: Autosomal Recessive
AD: Autosomal Dominant
XLR: X-linked Recessive
1CADD: Combined Annotation Dependent Depletion
OMIM: Online Mendelian Inheritance in Man
DEGs: Differentially Expressed Genes
SIFT: Sorting Intolerant from Tolerant
mCSM: Mutation Cutoff Scanning Matrix
DUET: Dual Enrichment Testing
PTMs: Protein Translational Modification
KEGG: Kyoto Encyclopedia of Genes and Genomes
RMSD: Root Mean Square Deviation
RMSF: Root Mean Square Fluctuation
Rg: Radius of Gyration
SAS: Solvent-Accessible Surface Area

## 6. Acknowledgement

We gratefully acknowledge the Indian Council of Medical Research (ICMR) for providing the Senior Research Fellowship (SRF) to PR and CD.

## 7. Author Contribution

**PR** conceived the research idea, conducted experiments, performed bioinformatics and data analysis, and wrote the manuscript. **CD** contributed to data analysis. **NV** was responsible for identifying CTA patients and conducting OPG analysis. **RB** and **VKS** carried out clinical investigations. **PD** co-conceived the research idea and approval of final draft.

## 8. Declaration

### Consent to participate

All subjects participating in this study were fully informed about the purpose, process, potential risks and benefits of the study and voluntarily signed a written informed consent form.

### Human ethics

**Ref No: I.Sc./ECM-XII/2021-2024:** The research protocol received approval from the Ethics Committee of the Institute of Science, Banaras Hindu University, UP, Varanasi, India.

### Data availability

The data supporting this study can be made available upon request.

