## Supplementary file 1 for "Whole Exome Sequencing Uncovers Key Genetic Variants in Congenital Tooth Agenesis: An Integrative Omics Approach"

Table 1: Relative Entropy Scores Computed by remuRNA

| Gene | Coding sequence change | Protein change | Relative Entropy  H(wt:mu) | MFE(mu) | MFE(wt) | dMFE |
| --- | --- | --- | --- | --- | --- | --- |
| *WNT10A* | c.433G>A | p.Val145Met | 0.355 | -13.2 | -12.7 | 0.5 |
| *WNT10A* | c.403G>T | A135S | 3.409 | -8.6 | -11.3 | -2.7 |
| *EDA* | c.467G>A | R156H | 2.248 | -4.1 | -4.1 | 0 |
| *TSPEAR* | c.656T>C | p.Leu219Pro | 1.392 | -12 | -10.6 | 1.4 |

The table presents computed structural features for wildtype (WT) and mutant (mu) RNAs, including relative entropy (H(WT:mu)), minimum free energy (MFE) for mutant RNA (MFE(mu)), MFE for wildtype RNA (MFE(WT)), and the difference in MFE between mutant and wildtype RNA (dMFE). The relative entropy, calculated by remuRNA, measures the localized structural differences between the Boltzmann ensembles of wildtype and mutant RNAs around the single nucleotide change of interest, thereby reducing the effects of sequence length. A higher relative entropy value indicates a stronger structural impact of the variant on RNA.

**Table2: Molecular Simulation Analysis Parameters of Gene Variants.** The table presents the molecular simulation analysis parameters for different variants of multiple genes.

|  | **EDA_WT** | **EDA_R156H** | **PAX9_WT** | **PAX9_Q145** | **TSPEAR_WT** | **TSPEAR_419fs150** | **TSPEAR+ Compound_ Hetro** | **WNT10A_WT** | **WNT10A_V145M** | **WNT10A_A135S** |
| --- | --- | --- | --- | --- | --- | --- | --- | --- | --- | --- |
| **RMSD** | 2.37 | 3.073 | 1.584 | 1.373 | 1.276 | 1.60 | 1.911 | 0.128 | 0.595 | 0.519 |
| **RMSF** | 0.690 | 1.106 | 0.415 | 0.343 | 0.366 | 0.366 | 0.675 | 0.239 | 0.227 | 0.264 |
| **SAS** | 18.779 | 124.049 | 72.57 | 86.325 | 262.05 | 190.50 | 201.06 | 137.268 | 135.551 | 135.563 |
| **H-Bonds** | 837.97 | 821.421 | 659.27 | 767.02 | 421.84 | 251.922 | 250.722 | 174.89 | 173.52 | 186.257 |
| **Rg** | 1.982 | 2.244 | 1.411 | 1.372 | 2.121 | 1.988 | 2.349 | 1.86 | 1.835 | 1.841 |

**Note:** The table presents molecular dynamics parameters for wild-type (WT) and mutant forms of EDA, PAX9, TSPEAR, and WNT10A. RMSD indicates structural deviation, RMSF shows residue flexibility, SAS reflects exposed surface area, H-Bonds represent structural stability, and Rg indicates protein compactness during simulations.

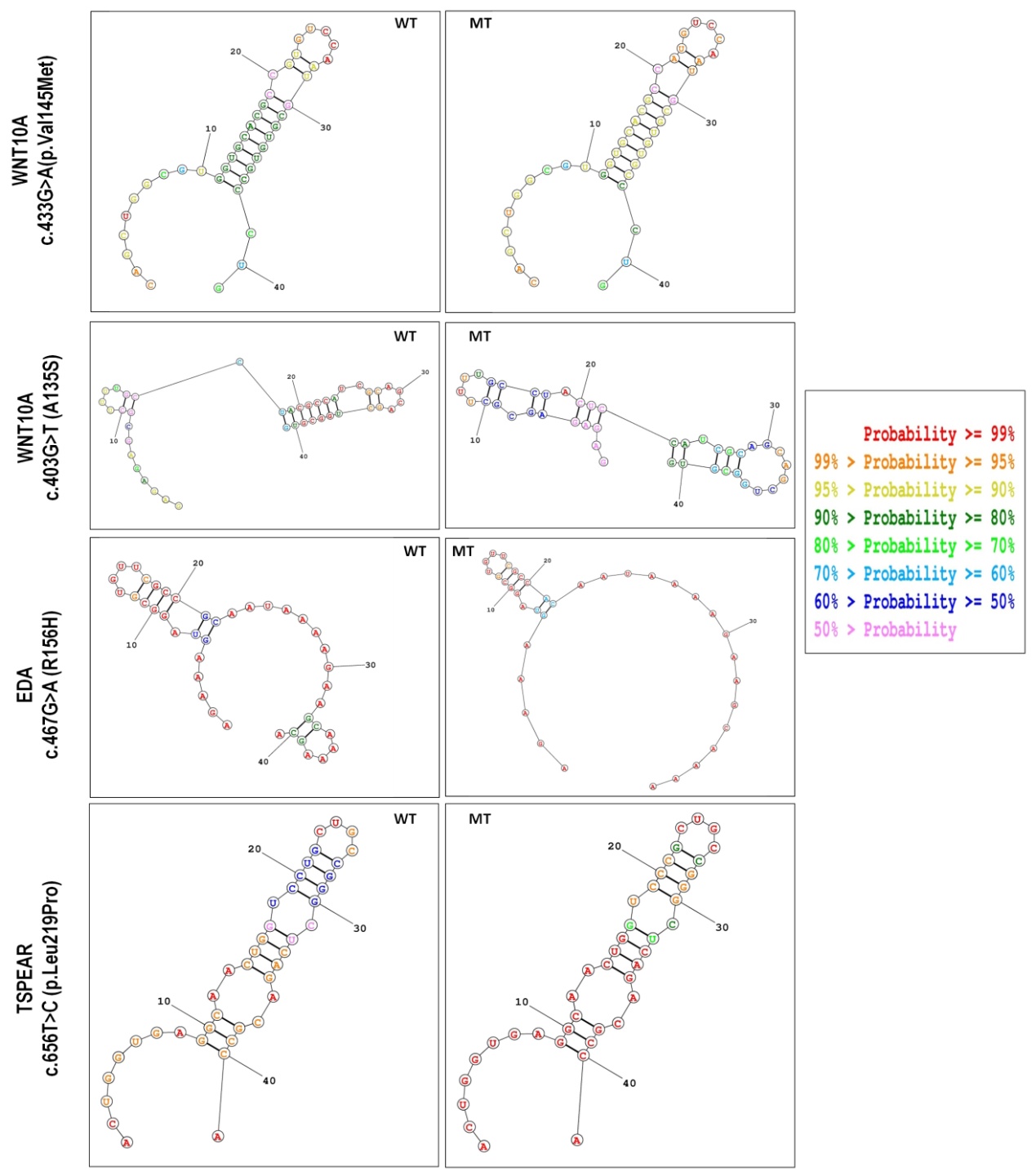

**Figure 1: Comparison of RNA Secondary Structures between Wild-Type (WT) and Mutant (MT) Snippets for Missense Variants of Interest.** This figure displays the predicted RNA secondary structures for wild-type (WT) and mutant (MT) snippets corresponding to each missense variants using the RNA Structure Web server. On the left side of the figure are the secondary structures of each WT RNA, while on the right side are the structure of the mutant RNA. The nucleotide change is at the 21st position in each structure. Various colors are utilized to represent different base pairing probabilities within the structures.

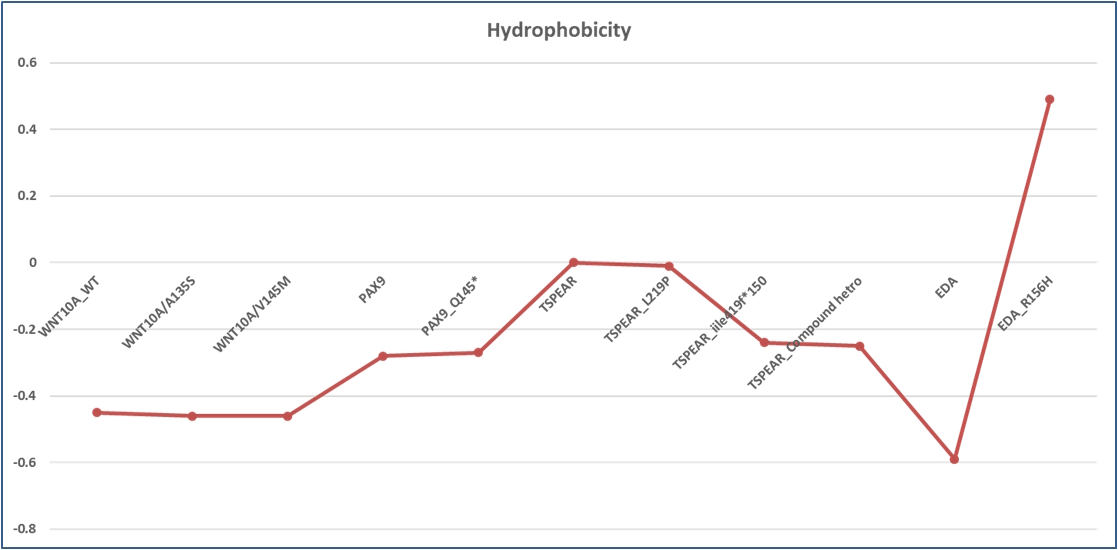

**Figure2: Hydrophobicity analysis of WNT10A, EDA, PAX9, and TSPEAR variants.** The figure depicts the hydrophobicity scores for each variant, calculated using a suitable hydrophobicity scale. The x-axis lists the variants, and the y-axis represents the hydrophobicity score. A positive score indicates hydrophobicity, while a negative score indicates hydrophilicity. The results highlight potential differences in the hydrophobicity profiles of the variants, which may influence their protein-protein interactions, stability, and localization.
